# APPLICATION OF THE ACMG/AMP FRAMEWORK TO CAPTURE EVIDENCE RELEVANT TO PREDICTED AND OBSERVED IMPACT ON SPLICING: RECOMMENDATIONS FROM THE CLINGEN SVI SPLICING SUBGROUP

**DOI:** 10.1101/2023.02.24.23286431

**Authors:** Logan C. Walker, Miguel de la Hoya, George A.R. Wiggins, Amanda Lindy, Lisa M. Vincent, Michael T Parsons, Daffodil M Canson, Dana Bis-Brewer, Ashley Cass, Alexander Tchourbanov, Heather Zimmermann, Alicia B Byrne, Tina Pesaran, Rachid Karam, Steven Harrison, Amanda B Spurdle

## Abstract

The American College of Medical Genetics and Genomics (ACMG) and the Association for Molecular Pathology (AMP) framework for classifying variants uses six evidence categories related to the splicing potential of variants: PVS1 (null variant in a gene where loss-of-function is the mechanism of disease), PS3 (functional assays show damaging effect on splicing), PP3 (computational evidence supports a splicing effect), BS3 (functional assays show no damaging effect on splicing), BP4 (computational evidence suggests no splicing impact), and BP7 (silent change with no predicted impact on splicing). However, the lack of guidance on how to apply such codes has contributed to variation in the specifications developed by different Clinical Genome Resource (ClinGen) Variant Curation Expert Panels. The ClinGen Sequence Variant Interpretation (SVI) Splicing Subgroup was established to refine recommendations for applying ACMG/AMP codes relating to splicing data and computational predictions. Our study utilised empirically derived splicing evidence to: 1) determine the evidence weighting of splicing-related data and appropriate criteria code selection for general use, 2) outline a process for integrating splicing-related considerations when developing a gene-specific PVS1 decision tree, and 3) exemplify methodology to calibrate bioinformatic splice prediction tools. We propose repurposing of the PVS1_Strength code to capture splicing assay data that provide experimental evidence for variants resulting in RNA transcript(s) with loss of function. Conversely BP7 may be used to capture RNA results demonstrating no impact on splicing for both intronic and synonymous variants, and for missense variants if protein functional impact has been excluded. Furthermore, we propose that the PS3 and BS3 codes are applied only for well-established assays that measure functional impact that is not directly captured by RNA splicing assays. We recommend the application of PS1 based on similarity of predicted RNA splicing effects for a variant under assessment in comparison to a known Pathogenic variant. The recommendations and approaches for consideration and evaluation of RNA assay evidence described aim to help standardise variant pathogenicity classification processes and result in greater consistency when interpreting splicing-based evidence.

## INTRODUCTION

In 2015, the American College of Medical Genetics and Genomics (ACMG) and the Association for Molecular Pathology (AMP) reported a framework for classifying variants using multiple evidence categories.^1^ Since this report, the Clinical Genome Resource (ClinGen) Sequence Variant Interpretation (SVI) Working Group have developed further guidance to applying different codes, for example PVS1 for loss of function variants,^2^ PS3/BS3 for variants impacting gene function, ^3^ and the stand-alone rule BA1 based on variant allele frequency.^4^

Although the ACMG/AMP guidelines recommend assessing whether gene variants could have an impact on natural RNA splicing, there is no standardised approach to interpreting this molecular information. Moreover, the complexities of the splicing process leads to challenges interpreting data generated from different computational predictions and laboratory assays. To date, the level of information in gene-specific ACMG/AMP guidelines provided by different ClinGen Variant Curation Expert Panels (VCEPs) for relevant codes (Table S1) has differed significantly, likely increasing uncertainty for diagnosticians. For example, when considering the functional assay codes (PS3/BS3) for splicing assays, the level of information provided by VCEPs ranges from no change to the original ACMG/AMP rules to detailed guidance for determining the appropriate strength of evidence based on assay type, assay outcome, and/or gene-specific variant location. The recommendations of Brnich et al for the application of functional PS3/BS3 criteria noted that splicing assays could be used to strengthen support for computational predictions for variants outside the canonical splice sites.^3^ However, there is a lack of guidance for applying and combining evidence codes based on splicing predictions, splicing assay data, and other functional data.

Bioinformatic tools that predict variant impact on splicing play a significant role in the assessment of variants of uncertain clinical significance. However, computational analysis of potential spliceogenic variants (defined as variants causing an altered mRNA transcript profile compared to control samples^5^) is challenging for variant curators due to the increasing number of tools available, each with bespoke settings that may not have been clinically validated. For the application of the PP3/BP4 codes, VCEPs differ significantly in their rules defining the level of computational evidence required to indicate a deleterious splicing effect (Table S1). Such differences include the number of splicing prediction tools used, the type of tool(s) to be used, and the thresholds to be applied for each. Indeed some VCEPS require multiple splice predictors to agree to apply codes, which may be inappropriate if the specified tools are designed to assess variant impact on different splicing motifs. Although a number of studies have compared the sensitivity and specificity of different splicing prediction tools, ^6–11^ there has been a lack of guidance on how to apply existing and future tools within the ACMG/AMP framework. Moreover, as there is such a high correlation between presence of variation at specific motif positions and likelihood to alter splicing, use of both position/prediction information and splicing assay data for interpretation might be considered redundant or overweighting.

Another two codes from the 2015 guidelines that are or may be used for variant interpretation relating to potential splicing impact are PVS1 (null variant in a loss of function gene) and BP7 (synonymous change with no predicted impact on splicing). Additionally, PS1 (same amino acid change as a previously established Pathogenic variant) could also be adapted for application to splicing-based evidence.

In response to groups seeking guidance on variant interpretation using splicing related evidence, the ClinGen SVI Working Group established the SVI Splicing Subgroup. In this manuscript, we detail evidence-based recommendations regarding the application of computational splicing prediction tools and in vitro splicing assays using a refined version of the ACMG/AMP sequence variant interpretation framework. We also provide recommendations on how to develop a gene-specific PVS1 decision tree and how to combine evidence codes for splicing and protein function derived from computational predictions and experimental assay data.

## MATERIAL AND METHODS

### Establishment of the ClinGen Sequence Variant Interpretation Splicing Subgroup

The remit of the ClinGen SVI Splicing Subgroup was to refine recommendations for applying ACMG/AMP codes relating to splicing data and computational predictions. ACMG/AMP criteria specifically evaluated for application (or adaptation) to impact on splicing included PVS1, PS1, PS3, PM5, PP3, BS3, BP4, and BP7. The goals of the subgroup were to: 1) determine the appropriate strength of evidence that can be applied to experimental splicing data for variant interpretation and appropriate criteria code selection for general use; 2) develop a framework for applying gene-specific PVS1 criteria; and 3) provide guidance on approaches to select and implement computational tools. Throughout this process we exchanged knowledge with several ClinGen VCEPs as they applied or developed ACMG/AMP specifications for different genes, and incorporated relevant information from these parallel efforts into our recommendations.

A review of the 53 established ClinGen VCEPs revealed that 23 panels had publicly available classification rules for one or more disease associated genes (Table S1). VCEP specifications reviewed and referred to in the text are shown in Table S1, and were as documented at 12 February 2023; please refer to the ClinGen Criteria Specification Repository for the most up-to-date versions of VCEP specifications (https://cspec.genome.network/cspec/ui/svi).

### Dataset of annotated canonical splice site variants identified in the diagnostic setting

A dataset comprising all individuals from a clinical (disease-affected) cohort referred to GeneDx (https://www.genedx.com/) for diagnostic testing over a 2 year time period was queried to extract information for all canonical splice site variants associated with known clinically relevant transcripts. A total of 1,447 canonical splice site variants were identified in 1,043 genes currently associated with disease through a loss of function mechanism (Table S6). SpliceAI scores were computed for the variants using default settings. The position of any predicted donor or acceptor site loss was manually verified to occur at the actual natural site for the transcript being evaluated. Predicted donor or acceptor gains were also checked to ensure they occurred within the appropriate sequence context. Variant classification had previously been determined following GeneDx clinical protocols in line with application of ACMG/AMP criteria.^1^ These protocols included review of in silico prediction data (to assess predicted impact on transcript/s and PVS1 strength), published literature pertaining to the variant, and clinical data for the proband.

### Computational, splicing assay and functional datasets

Two large truth datasets were used to assess utility of computational prediction tools. The first dataset contained cell survival results (phenotypes: loss of function, intermediate function and functional) from 414 *BRCA1* variants^12^ supplemented with computational splicing prediction results (this study). While the cell survival functional assay did not provide specific results about splicing events, the results were used to infer whether variants were functionally benign or pathogenic, which for intronic variants was presumed to be due to impact via RNA effects. The analysis dataset included information for variants at the donor and acceptor region (3 nucleotides exonic (synonymous substitutions only) and 8 nucleotides intronic).

The second dataset contained in vitro splicing assay and computational splicing prediction results across a range of disease susceptibility genes that have been curated from the literature: 1,008 *BRCA1/BRCA2* variants, 659 mismatch repair gene variants (*MLH1, MSH2, MSH6*, *PMS2*),^13^ 284 *NF1* variants,^6^ and 1,070 *POU1F1* variants. ^14^ The majority of these variants were patient-identified. The *POU1F1* dataset differed in that it represented results from a high-throughput assay designed to test the effect of variants on upregulation of a minor isoform with transcriptional repressor activity. Of the 3,021 variants collated for this study, 767 were reported to be spliceogenic (associated with one or more aberrant splicing events). All variant data are provided in Table S2 and available for download from a web tool we developed to facilitate calculation of likelihood ratios for calibration (https://gwiggins.shinyapps.io/lr_shiny/). Based on a conservative interpretation of the position weight matrix plot for U2-type introns (the most prevalent intron type),^15^ the following variant categories were created for the main analysis, based on variant position relative to the canonical splice sites: 1) Canonical splice site (±1/2 intronic nucleotide positions); 2) Standard splice region (Donor site motif - last 3 bases of the exon and 6 nucleotides of intronic sequence adjacent to the exon; and Acceptor site motif - first base of the exon and 20 nucleotides upstream from the exon boundary); 3) “Other” - intronic or exonic nucleotide positions outside canonical splice site and standard splice region. Additionally, sensitivity and specificity was assessed for variants within a minimal splice region (Donor site motif - last 3 bases of the exon and 6 nucleotides of intronic sequence adjacent to the exon; and Acceptor site motif - first base of the exon and 3 nucleotides upstream from the exon boundary), and then separately for intronic and exonic nucleotide positions outside of canonical splice sites and the minimal splice region.

### Splicing prediction analysis

The deep learning–based splice variant software SpliceAI^16^ (https://github.com/Illumina/SpliceAI - Version 1.3.1) was used to predict the effect on splicing for each variant. We used the maximum raw Δ score, defined as the maximum probability of altered splicing across four output probabilities (loss of acceptor or donor sites, gain of acceptor or donor sites), and the maximum distance of 10,000 nucleotides (±4,999 nucleotides from the variant of interest). The performance of SpliceAI was compared with 10 other splicing prediction tools (listed in Table S3) using the *BRCA1* variant survival functional dataset from Findlay et al,^12^ which consisted of single nucleotide variants at or near 13 *BRCA1* exons that encode functionally critical domains. Each variant had been assessed and categorized as functional (FUNC) or non-functional (Loss of Function [LOF]) based on the outcome of a survival assay using HAP1 cells. Variants with intermediate function were excluded from the analysis. A total of 414 variants (n=312 FUNC and n=102 LOF) were used for the evaluation of prediction tools.

The relative performance of the splicing prediction tools was evaluated using Receiver Operating Characteristic (ROC) curve analysis. Specificity was defined as the total true negatives out of all negatives (true negatives + false positives). Sensitivity was defined as the total true positives detected out of all positives (true positives + false negatives). The prediction tool SpliceScan II was modified to include: (1) a Bayesian splice site sensor,^17^ (2) evolutionary conservation, and (3) the Bayesian classification framework as introduced by Tavtigian et al to score splice site motifs based on both splicing biology and variant pathogenicity,^18^ the modified framework is termed SplicScan III. Only SpliceAI scores were generated for the literature-curated dataset of variants.

### Calibration of code weights based on odds of spliceogenicity

Likelihood ratios (LRs) for prediction of spliceogenicity - as a measure of inferred pathogenicity - were estimated for different SpliceAI categories using our custom-built web tool (https://gwiggins.shinyapps.io/lr_shiny/; See Table S4 for a worked example). An iterative approach was used to select the best cut-off that minimised apparent false-negative and false-positive predictions, and the overall proportion of variants within the uninformative zone. Sensitivity analysis was conducted to assess robustness of LR estimates for a narrower band of scores approaching the selected score cut-offs. Designation of LRs to ACMG/AMP rule code strengths were based on LR ranges recently proposed as consistent with ACMG/AMP qualitative rule strengths for future classification in a Bayesian framework.^18^ The benign (or non-spliceogenic) category intervals were calculated as inverse odds to the pathogenic (or spliceogenic) category intervals. These odds ranges assume a global prior probability of pathogenicity of 0.10. Bayesian pathogenicity criteria thresholds are shown in Table S5.

### Designation of donor and acceptor motif ranges for bioinformatic code application

Donor and acceptor motif ranges are relevant for application of codes PS1 and BP7. The vast majority of introns (>98%) are recognised by highly conserved dinucleotides at the 5′ boundary (GT) and 3′ boundary (AG).^19; 20^ Intron categories may variously be designated by the boundary dinucleotide sequence, spliceosome (likely) excising the intron (U2-type spliceosomes for most GT-AG introns, U12-type spliceosomes for most AT-AC introns, with some exceptions), and/or by comparing position weight matrices for surrounding sequence.^20; 21^ The standard splice region designated for donor and acceptor site motifs consisted of the last 3 bases of the exon and 6 nucleotides of intronic sequence adjacent to the exon, and the first base of the exon and 20 nucleotides upstream from the exon boundary, respectively. This motif range may be altered for more detailed variant-specific analysis, if considered appropriate e.g. for the much rarer U12-type introns, conservation maps suggest that the donor site motif should include only the last base of the exon and be extended to 9 nucleotides of intronic sequence adjacent to the exon. Additionally, the minimal splice region included donor region as defined above, with the acceptor site motif designated as the first base of the exon and 3 nucleotides upstream from the exon boundary).

## RESULTS & DISCUSSION

### Review of existing Variant Curation Expert Panel guidelines

The ACMG/AMP guidelines for splicing-related results provide a framework for classifying variants in disease susceptibility genes, most relevant to those genes where loss of function is the mechanism of pathogenesis. Adaptations of the original guidelines reported by Richards et al^1^ have been described by ClinGen VCEPs, which enable curators to apply modifications to the relevant strength of each evidence type and gene-specific considerations. A review of the 53 established ClinGen VCEPs revealed that 23 panels had publicly available classification rules (Table S1). We also included pilot rules from the ENIGMA BRCA1 and BRCA2 VCEP, the Hereditary Breast, Ovarian and Pancreatic Cancer VCEP (PALB2) and InSiGHT Hereditary Colorectal Cancer/Polyposis VCEP (Mismatch repair genes), which developed their specifications for splicing-related codes in parallel with the activities of the ClinGen SVI Splicing Subgroup. In addition, code specifications and detailed advice from the Cancer Variant Interpretation Group UK,^22^ a major clinical network operating independently of ClinGen, were also reviewed. Comparisons of these accumulated specifications demonstrate between-panel variation in application of ACMG/AMP codes (and code strengths) pertaining to splicing-related evidence.

For interpretation of variants impacting the canonical splice site dinucleotides, rules from six ClinGen VCEPs and the Cancer Variant Interpretation Group UK use the original version of the PVS1 decision tree published by Abou Tayoun et al (Abou Tayoun et al., 2018) to assign a final weight of evidence. As detailed in Table S1, 15 VCEPs developed gene-specific PVS1 guidelines by modifying the branches of the original decision tree. In addition, some VCEPs (e.g. CDH1, ENIGMA, Familial Hypercholesterolemia, Hereditary Breast Ovarian and Pancreatic Cancer, InSiGHT Hereditary Colorectal Cancer/Polyposis, Lysosomal Storage Disorders, and Rett and Angelman-like Disorders) and the Cancer Variant Interpretation Group UK provide scope for adapting other codes that address splicing, such as PS3/BS3, based on the level of evidence strength (supporting>moderate>strong>very strong). Importantly, there is marked variability between the different VCEP guidelines for applying ACMG/AMP splicing-related evidence types, variability which cannot be solely explained by gene-related and disease-specific factors. For example, guidelines on the number of splicing prediction tools required to warrant the PP3 code (computational evidence to support splicing) ranged from one to three tools. Notably, a limited number of VCEPs incorporated the interpretation of splicing prediction data into modifications of the missense-based codes PS1 (DICER1 and miRNA-Processing Gene, ENIGMA, Hearing Loss, Hereditary Breast Ovarian and Pancreatic Cancer, InSiGHT Hereditary Colorectal Cancer/Polyposis, Monogenic Diabetes, and PTEN) or PM5 (CDH1 and InSiGHT Hereditary Colorectal Cancer/Polyposis).

The following sections outline processes for establishing a gene-specific framework to derive strength for ACMG/AMP splicing-related evidence types for use in clinical variant interpretation.

### Development and application of a gene-specific PVS1 decision tree for splice site variants

#### Assessing relevance of the PVS1 code for a given gene

PVS1 is a predictive code for (presumed) loss-of-function (LoF) variants, including nonsense, frameshift, canonical splice site, single or multi-exon deletions/duplications, and the initiation codon.^1^ This code is applicable to a large proportion of genes where LoF is a known mechanism of disease. Establishing LoF as a disease mechanism can be subjective; however, several helpful resources to assess LoF include the ClinGen haploinsufficiency (HI) score,^23^ the probability of LoF intolerance score (pLI ^24^), and/or the “loss-of-function observed/expected upper bound fraction (LOEUF ^25^). HI score availability is limited as it is based on manual curation of evidence with an output divided into six tiers, where a score of “3” indicates “Sufficient evidence suggesting dosage sensitivity is associated with clinical phenotype” (curations available at: https://search.clinicalgenome.org/kb/gene-dosage). Comparatively, pLI scores are computationally derived, and measure the intolerance of a given gene to LoF variants in the general population; scores are available in gnomAD ((https://gnomad.broadinstitute.org/), with a pLI > 0.9 suggesting a significantly lower than expected rate of LoFs in the gene.^24^ As the pLI value can be significantly influenced by sample size (i.e. a gene with low expected allele counts across the gene could not have a high pLI), LOEUF (available with gnomAD v2.1 release) is now recommended over pLI for LoF intolerance assessments; a LOEUF threshold of <0.35 suggests LoF intolerance that is similar to the pLI > 0.9 threshold. It is important to note that both pLI and LOEUF predictions are dependent on transcript selection and measure intolerance relative to reproductive fitness, thus age of onset and disease severity must be considered when assessing whether LoF variants are expected to be observed in population datasets. If a gene can predispose to disease via both LoF and gain-of-function (GoF) mechanisms, and a PVS1-eligible variant leads to a predicted (or experimentally observed) aberration expected to result in GoF, we recommend use of the PM4 code (described as “protein length changes due to in-frame deletions/insertions in a non-repeat region or stop-loss variants”) as opposed to PVS1, since PVS1 is specific to variants where LoF is the known disease mechanism.^26^

#### Construction of a gene-specific PVS1 decision tree

While most genetic alterations considered under the PVS1 code are *bona fide* LoF variants, it is critical to evaluate variants in the context of gene structure and expression, including the impact of the variant on alternative splicing, mRNA stability, and function of resultant protein products. For that reason, the ClinGen SVI previously developed recommendations for PVS1 application^2^ that specifically addressed four “rescue” mechanisms that may modulate the functional and clinical impact of assumed LoF variants: 1) premature termination codons (PTCs) at the 3’ end of the coding sequence leading to a shorter yet still functional protein; 2) stop-gains (nonsense and frameshift variants) or deletions located in or encompassing non-constitutive exons; 3) in-frame splicing or genomic deletions/duplications leading to a shorter or longer yet still functional protein; and 4) rescue of initiation codon variants by use of an alternative in-frame ATG.

With regard to canonical splice sites, the final PVS1 code strength depends on which of these four rescue mechanisms may be relevant after review of the predicted consequence/s of the splice site change. Here we highlight key considerations and a process for developing a gene-specific PVS1 decision tree.

##### i) Characterising expression and structure of the reference transcript

Differences in transcript structure resulting from alternative splicing can influence cellular function and contribute to disease. Furthermore, outcomes of abnormal splicing are typically varied with respect to transcript levels and the resulting protein-reading frame. Accurate variant interpretation for PVS1 requires scientific knowledge of the structure and function of transcripts expressed by each gene being examined. As noted previously by Richards et al,^1^ a reference transcript for each gene should be identified, utilised and reported when describing variants being investigated. To promote consistency for clinical relevance, the National Center for Biotechnology Information (NCBI) and the European Molecular Biology Laboratories-European Bioinformatics Institute (EMBL-EBI) collaborated on the release of reference transcripts through <u>M</u>atched <u>A</u>nnotation from <u>N</u>CBI and <u>E</u>MBL-EBI (MANE - https://www.ncbi.nlm.nih.gov/refseq/MANE/).^27^ One transcript for each protein-coding locus deemed the MANE Select is annotated as the “default” reference across genomic resources, including UCSC Browser (GRCh38; https://genome.ucsc.edu/), gnomAD v3.1.1, and ClinVar. While MANE Select is supported by experimental data, the single reference transcript does not necessarily capture biological complexity of disease causation. A second initiative, MANE Plus Clinical, captures secondary transcripts that contain Pathogenic variants not found in the MANE Select transcripts, or with more impactful effects (exonic versus intronic).

##### ii) Mapping regions critical to protein function

Interpreting splicing effects of each gene transcript requires knowledge of regions critical to protein function. Identifying protein functional domains utilizes homology-based predictive analyses (e.g. InterPro, Pfam), supportive functional data (e.g. deletion of the protein domain has a measurable impact on a functional assay), and/or structural data (e.g. crystal structure of a protein complex shows residues directly interacting with a critical partner). Prior knowledge of Pathogenic missense variants may also highlight regions critical to protein function and clinical relevance. However, the overall quality and granularity of the protein mapping depends on the level of information available for each specific gene. We propose updates to the Abou Tayoun et al PVS1 decision tree^2^ (see Table 1) including assigning very strong evidence of pathogenicity to in-frame RNA skipping events encompassing undisputed clinically relevant residues. Note, the final PVS1 weighting may be reduced depending on the structural features of the critical region and size/location of the in-frame alteration (See Supplemental Information, Box S1: Biologically relevant transcripts and the rescue transcript model). Further, final PVS1 weighting may be strengthened when variant/s at complementary splice sites (i.e. the acceptor and donor sites for the same exon) with the same predictions (e.g. in-frame exon skipping) have been classified as Pathogenic. (Note, the term classified is defined here as variant curation following ACMG/AMP guidelines, and preferably following VCEP gene-specific recommendations if available).

**Table 1.** Splicing-relevant modifications to the PVS1 decision tree of Abou Tayoun et al (2018)^2^

| Original Abou Tayoun et al PVS1 decision tree | New recommendations | Example in Figure 1 |
| --- | --- | --- |
| Splicing alterations at 5' (or 3') UTRs are not specifically addressed | Annotate as PVS1_N/A unless the splicing outcome is predicted to impact protein or elements proven critical for function and/or expression. | A |
| Apply PVS1_Moderate if there are no alternative start codons in the transcript set for a gene, for a start loss variant if one or more Pathogenic variant(s) have been reported 5' of the next downstream putative in-frame start codon. | Consider upweighting based on the frame of alternative start sites, their location relative to known clinically important functional domains, and/or location of Pathogenic variants 3' to the native start site and 5' to the next putative in-frame alternative start site. | B |
| Apply PVS1_Strong for single- to multi-exon deletions that preserve reading frame, and truncated/alterd region is critical to protein function. Relevant domain indicated by experimental evidence proving a critical role of the domain and/or presence of non-truncating pathogenic variants in the region. | Consider upweighting to PVS1 if the in-frame deletion removes a protein region critical to function e.g. Pathogenic missense variant has been identified in that protein domain. | F |
| If >10% of the protein is removed, apply PVS1_Strong, regardless of whether the region is critical to protein function. | Consider upweighting to PVS1 if the truncated protein region is critical to function e.g. Pathogenic missense variant has been identified in that protein domain or complementary splice site Pathogenic variation. | H |

##### iii) Building reference gene-specific splicing catalogues

Annotating comprehensive catalogues of naturally occurring splicing events that occur across different tissue types are critical for building reference datasets to guide the application of gene-specific ACMG/AMP codes, and indeed have been (or are being) used by several VCEPs.^28–36^ The profile can be derived from publicly available curated databases (e.g., GENCODE Basic), in-house analysis of RNA-seq repositories, and/or from a dedicated experimental approach. The quantity and quality of data might be variable for different genes, thus affecting the body of knowledge available for gene-specific adaptations of the PVS1 decision tree. Most technologies (conventional RT-PCR approaches, targeted and whole-transcriptome RNA-seq) analyse partial transcript sequences, providing information on alternative splicing events rather than on the complete exon structure of alternative mRNA isoforms. Long-read sequencing technologies may also be utilized to resolve complex exon structures of full-length transcripts without the need to bioinformatically reconstruct sequences.^37^ Employing such an approach has previously shown that some alternative transcripts contain multiple alternative splicing events,^38; 39^ which has potential implications for determining the coding frame, interpreting the clinical significance of spliceogenic variants, and identifying naturally occurring candidate rescue transcripts (see next section). Comprehensive long-read RNA-seq data is currently lacking for many disease susceptibility genes. Furthermore, these alternative splicing data also provide a useful diagnostic resource for the design and interpretation of in vitro splicing assays.

##### iv) Identifying naturally occurring candidate rescue transcripts

Some consensus splice site variants impact non-constitutive exons in alternatively spliced transcripts. Splicing of naturally occurring transcript(s) *excluding* that exon will not be impacted, and if predicted to encode a functional protein, are considered to be candidate rescue transcript(s). The same rescue model is pertinent to the classification of stop-gain variants (nonsense and frameshift variants) in that exon and deletions of that exon since the relevant variants would be absent from rescue transcripts. (See Box S1 and Figures S1-3 for further description of the rescue transcript model and identification of transcripts with potential to contribute to a rescue mechanism.)

The initial step to identify candidate rescue transcripts is to review alternative transcripts against a gene-specific map of critical protein domains; transcripts preserving the reading frame and coding for critical protein domains are candidate rescue transcripts. We recommend annotating physiological alternative splicing events as candidate rescue transcripts even if tissue-specific expression is uncertain or unknown (e.g. due to lack of data). The appropriate baseline expression threshold for designating a candidate rescue transcripts will likely be gene-specific and ideally should be based on experimental data. We anticipate such data will be scarce for many clinically relevant genes and propose 10% of the overall gene expression as an operational threshold (Figure S3). We propose use of PVS1_N/A for variants for which there is a plausible rescue model, based on observation of naturally occurring alternative spliced transcripts.

##### v) Recommendations to improve gene-specific PVS1 decision trees

The main role of bioinformatic predictions for splice site variants is to determine if the expected predominant splicing alteration will be in-frame or out-of-frame. Thus, we recommend use of a tool that can predict not only spliceogenicity, but also the nature of the transcript produced by the splicing alteration (i.e. exon skipping and/or use of a cryptic/de novo site and/or intron retention). To further add to recommendations from the original PVS1 decision tree publication, we recommend the following generic issues to be considered:

- Bioinformatic scores to invoke abrogation of native canonical GT donor sites are likely to differ from those for native non-canonical sites.
- Some alterations at native splice sites are not predicted to alter splicing and should not be assigned a PVS1 evidence strength (PVS1_N/A). For example, a subset of IVS+2T>C variants at a canonical GT donor site will result in a functional GC donor site, and the bioinformatic score for a native GC site may be improved by a IVS+2C>T variant.
- The range of evidence strengths applicable for predicted in-frame alterations could be increased if justified by functional and/or clinical evidence. For example, substituting the generic 10% protein size threshold with protein specific thresholds based on protein structure considerations if available from the literature, or knowledge of a Pathogenic missense variant located within a functional domain. This update will affect GT-AG variants predicted to cause in-frame alterations (as noted above, this may include complementary acceptor and donor sites for the same exon), but also other in-frame alterations such as exon deletions. A gene-specific example can be seen in the PVS1 decision tree for *ATM* developed by the Hereditary Breast, Ovarian and Pancreatic Cancer VCEP (https://clinicalgenome.org/site/assets/files/7451/clingen_hbop_acmg_specifications_atm_v1_1.pdf).

Given these considerations, our recommended modifications to the original decision tree published by Abou Tayoun et al (2018) are summarized in Table 1 in conjunction with examples of PVS1 strength modifications for canonical splice variant effects within different gene-specific contexts (Figure 1).

**Figure 1.**
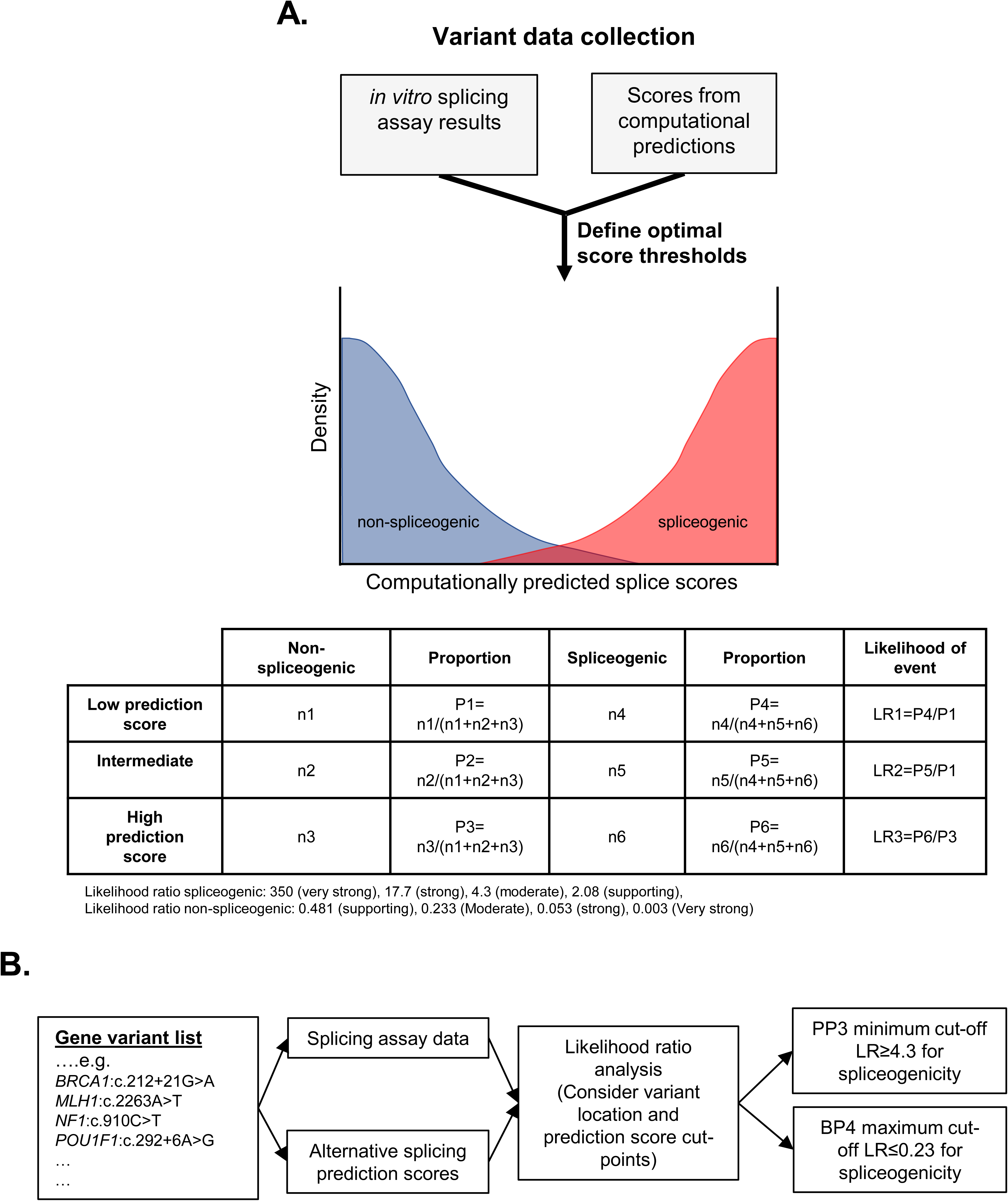
Schematic demonstrating assignment of gene-specific codes to canonical splice sites based on a modified version of the PVS1 framework proposed by Abou Tayoun et al., 2018.^2^ It is important to note that each PVS1 assigned weight may be reduced if there is evidence of potential rescue mechanisms. For example, skipping of either exon 4 or 7 may lead to a protein that retains partial function. Annotating gene-specific lists of naturally occurring splicing events can provide greater evidence of potential ‘rescue’ isoforms. Also see Supplemental Information, Box S1, Biologically relevant transcripts and the rescue transcript model.

### Re-purposing of the PVS1 and BP7 codes to capture observed *in vitro* splicing data irrespective of variant location

To date, application of the PVS1 decision tree for splicing has focused on canonical ±1,2 splice site variants. We raise two points for consideration. The first is that variation at the canonical dinucleotides has such high probability to impact splicing that confirmation of variant effect by RNA assay data is generally unlikely to alter the starting PVS1 weight. The second point for consideration is that - if a variant located outside of the canonical dinucleotides is proven to result in an aberrant splicing profile that is interpretable via the PVS1 decision process - it can be assumed to have clinical impact equivalent to that of a canonical dinucleotide splice site variant assigned a PVS1 code (either predictive or based on observed experimental data) using the same PVS1 decision process. We note that these considerations are made assuming curation of variants in the context of Mendelian disease, the baseline assumption for the ACMG/AMP guidelines as originally published. For further guidance on interpretation of splicing profiles, see section below “Adaptive weighting of evidence based on splicing assay type, design and complexity of transcript profile”.

#### High correlation of splice site PVS1 weights with clinical findings and use of the PVS1 code to capture splice assay results

Experimental confirmation of a splicing event does not necessarily increase confidence in the clinical significance of that event. For example, a variant at the canonical dinucleotide position that is predicted to lead to an in-frame splicing event could be down weighted (PVS1_Strong or PVS1_Moderate), dependent on knowledge of functional relevance and/or the extent of protein lost (Figure 1). Confirmation of such an in-frame splicing event would not change confidence in the clinical significance of that event, and use of PS3 for results from splicing assays in addition to the relevant downweighted PVS1 code would overweight the evidence towards pathogenicity. For canonical splice site variants, splicing assay data has most value in resolving impact where predictions are less confident (e.g. +2T>C variants, ^40^) or may suggest multiple possible transcripts; establishing impact and weighting where multi-exon skipping (out of scope of predictions) is suspected. Where variant predicted impact on transcript profile differs from transcript/s identified using RNA assays, particularly in terms of designated weight, this information should be used to upgrade or downgrade the PVS1 code weight for that splice site in the context of a gene-specific decision tree.

Indeed, a review of clinical laboratory data on canonical splice site variants identified in 1043 genes currently associated with disease due to loss of function (listed in Table S6) supports the a priori assumption that - while the vast majority of such variants are disease-causing - additional curation is needed to refine PVS1 weighting. Of 3400 total canonical splice site variants in these 1043 genes, 3031 were internally classified as (Likely) Pathogenic (89%) and 347 as VUS (10%). Another 22 (0.6%) were classified as (Likely) Benign. Bioinformatically predicted impact was consistent with location of (Likely) Pathogenic variants at either acceptor or donor sites; i.e., acceptor site variants were enriched for high acceptor loss scores and donor site variants were enriched for high donor loss scores (Figure S4). As detailed in Table S7, classification for (Likely) Benign variants was largely based on population and/or benign clinical evidence. More detailed examination of (Likely) Benign variants revealed that five variants reported commonly in gnomAD were likely sequencing artefacts. Predicted splicing effect revealed that another 16 variants were unlikely to lead to functional impact due to: location in the 5’UTR, location in the last coding exon, predicted minimal effect on encoded protein length or read-through, predicted in-frame event, or no predicted effect on splicing. The remaining variant (*BRCA1* c.594-2A>C) is known to be Benign due to a rescue transcript mechanism (see Supplemental Information, Box S1). That is, having excluded sequencing artefacts and after consideration of rescue transcripts, these (Likely) Benign variants would not be assigned PVS1 based on the thorough application of a PVS1 decision tree (Figure 1).

Moreover, a total of 1670/3400 splice site variants were reported in publications; 404 of these had functional information available, of which 392 (97%) were classified as (Likely) Pathogenic. The remaining publications included cohort analyses, case studies, and review papers. When looking at all variants with publication data, 1576/1670 (94%) were (Likely) Pathogenic indicating that the vast majority of canonical splice variants that have been investigated are damaging. Closer review of the published variants revealed a large amount of evidence in favour of pathogenicity including: in silico (assessment of predicted null effect, presence at the same position as another Pathogenic variant, presence in clinical databases), laboratory data (functional assays) and/or clinical information (de novo status in affected individuals, in trans with a Pathogenic variant, identification in an individual with phenotypic fit, segregation with disease in families). To determine how often the classification was impacted by the application of in silico criteria alone, we evaluated how many (Likely) Pathogenic variants had clinical evidence (PS2, PM3, PM6, PP1, PP4) or laboratory evidence (PS3) versus only in silico evidence. For this comparison, PM2 was not considered as clinical evidence. PVS1 or PVS1_Strong was applied to 2894 (Likely) Pathogenic variants. At least one (and up to 7) clinical or laboratory criteria were applied to 1781/2894 (62%) variants in addition to PVS1 or PVS1_Strong. That is, only 38% relied on absence in controls, an indirect form of clinical data, to reach a Likely Pathogenic classification. These data further support the proposition that predicted disruption of the canonical splice site that meets at least a strong PVS1 code assignment leads to LoF.

To help distinguish when PVS1 is applied for canonical dinucleotide variants due to splicing assay data as opposed to predictions only, we recommend additional annotation to PVS1 application, such as PVS1_Strength (RNA). If the annotation of “RNA” to PVS1 is not available or possible in relevant curation systems, then curators should note that application of PVS1 was due to the presence of splicing assay data in both the explanation for criteria application as well as the overall variant evidence summary. Subsequently, it is recommended that the PS3 (or BS3) code is applied only for well-established assays assessing functional impact that is not directly captured by RNA splicing assays (e.g. in vitro assays that by design measure only effects on protein function or cellular assays that capture impact on protein function as well as on mRNA stability or processing).

#### Using PVS1 decision trees to apply PVS1 for confirmed spliceogenic variants outside of canonical splice sites

The second point for consideration relates to variants located outside of the canonical dinucleotides. If in vitro analysis of a variant under investigation results in an aberrant splicing profile that is interpretable via the PVS1 decision process, this can be assumed to have clinical impact equivalent to that of a canonical dinucleotide splice site variant assigned a PVS1 code (predictive or based on experimental data) using the same PVS1 decision tree. This allows a variant originally assigned only a supporting bioinformatic code (PP3 or perhaps even BP4 for poorly predicted splicing events such as exonic splicing enhancer alterations) to be assigned a PVS1 code based on the experimentally observed splicing event effect for LoF. That is, PP3 (or BP4) would be replaced with an appropriately weighted PVS1 code designation based on experimental data. As a result, variants observed to lead to the exact same splicing aberration (type and level) will receive the same experimentally-derived PVS1 weight irrespective of their location relative to the canonical dinucleotide positions.

#### Use of BP7 code to annotate absence of experimentally observed splicing impacts for silent substitution and intronic variants

As noted above, it is recommended that PS3 (or BS3) is applied only for well-established assays assessing functional impact that is not directly captured by RNA splicing assays. To distinguish when a silent substitution is confirmed to have no impact on splicing in vitro, we recommend upweighting BP7 with an additional annotation, namely BP7_Strong (RNA). Further, consistent with the extended use of BP7 to capture the low prior probability of pathogenicity for intronic variants with no predicted impact on splicing (see section “Application of the BP7 computational code”), we recommend that this same annotation may be used to capture *i*n vitro evidence of no splicing impact for intronic variants irrespective of position and predicted impact on splicing. We anticipate that this will be relevant almost exclusively for intronic variants outside of the native splice motifs, but would also be applicable in the unlikely event that a splice site variant does not impact splicing e.g. as has been reported for +2 C>T changes.^41^ If the annotation of BP7 to reflect RNA-assay is not possible in relevant curation systems, curators should note that application of BP7_Strong was due to the presence of splicing assay data in both the explanation for criteria application as well as overall variant evidence summary.

### Adaptive weighting of evidence based on splicing assay type, design and complexity of transcript profile

Many factors can influence results from splicing assays. Examples of considerations for both splicing assay design and the interpretation of results from splicing assays are listed in Table S8. These include RNA source, assay design, methodology and technology, and quantitative measurement of variant-impacted transcripts. The weight applied to PVS1 or BP7 codes based on experimental data should consider the confidence in the RNA findings having considered such factors. Where possible, gene-specific information should also be incorporated for determining most appropriate weights.

Conservatively, PVS1 and BP7 codes based on experimental RNA data may be considered applicable at full weight only for results from assays conducted on RNA from patient germline tissue samples (e.g. fresh blood, cultured lymphocytes, and lymphoblastoid cell lines, or relevant tissue types where gene expression is tissue-specific). Since expression of spliced transcripts can be tissue-specific,^42^ it is important that evidence for variant interpretation from splicing assays considers potential relationships between impact of spliceogenic variants and tissue-specific expression of any gene transcript.^43^ The Human Protein Atlas (https://www.proteinatlas.org/) provides a data resource of RNA expression from 40 non-diseased human tissue types and incorporates three large searchable datasets including: 1) Human Protein Atlas based on RNA-seq,^44^ including data from the Genotype-Tissue Expression (GTEx) project;^45^ and 2) the FANTOM consortium using cap analysis gene expression (CAGE) technology.^46^ Such datasets form the basis of categorising gene transcripts for splicing assays based on known expression levels and tissue distribution. Furthermore, they provide key information to consider how closely a splicing assay from the clinically accessible tissue (e.g. blood or fibroblast) reflects biological activity in the relevant disease tissue (e.g. breast or brain), and thereby assess the validity of the splicing assay. To remove the possibility of misinterpreting the clinical relevance of spliceogenic variants in patient samples, naturally occurring alternative splicing events must be established using control samples from matching tissue.

While assays using relevant patient material are generally considered preferable for assessing variant effect on RNA splicing, minigene constructs - as a hemizygous system - are useful in providing allele-specific quantitative measurements of exonic and particularly intronic variant impact on splicing. More recently, massively parallel reporter assays (MPRAs) have been applied to screen for spliceogenic effects of thousands of variants in a single experiment.^47^ Such data could be a rich source of information as a truth dataset for assessing the sensitivity and specificity of computational splicing prediction tools and also as a measure of splicing impact in the context of variant interpretation. Interpretation considerations of splicing results from these construct-based assays, include: 1) size constraints of constructs prevent analysis of very large exons and may exclude important regulatory sequences due to restrictions in extent of intronic sequence captured; 2) dependency on both the cell-type used for assays and the promoter used in the reporter construct; and 3) measurement of observed changes in splicing (e.g. using percent spliced in [ψ] from RNA-seq data) requires careful consideration when defining data thresholds for “impact on splicing” in different tissues. In light of these, a conservative approach would be to apply information from construct data alone at lower weight in the absence of calibration of an experimental system against clinical data for proven spliceogenic and non-spliceogenic variants.

Classification of incomplete (‘leaky’) splicing variants, that reduce but do not abolish normal splicing in the reference transcript, remain a challenge. The proportion of alternatively spliced gene transcript/s arising from a variant allele may have a significant impact on the severity of a patient’s clinical phenotype (variable expressivity), and/or be associated with reduced penetrance of the expected clinical phenotypes. It is therefore important to establish what proportion of alternatively spliced gene transcript arising from a variant allele will or will not confer pathogenicity for a given gene-disease relationship. For example, *BRCA1* variants resulting in 20-30% expression of a functional transcript should not be considered pathogenic for hereditary breast-ovarian cancer.^48^ By comparison, partial penetrance of autosomal recessive cystic fibrosis has been correlated with the level of the full-length *CFTR* transcripts in respiratory epithelial cells from spliceogenic variant carriers.^49^ Carriers with normal lung function and minimal/no lung disease demonstrate >25% of full-length CFTR transcripts is sufficient to maintain function.

Defining thresholds for incomplete splicing is complex and requires expert knowledge of the disease susceptibility gene. Quantitative assays, such as RNA-seq, are needed to ascertain the level of incomplete splicing for variant-carrier patients and controls. The extent of aberration induced by “leaky” variants may be evaluated by directly detecting allele-specific expression, for variants expressed in an exon or that activate a cryptic site within the intron, or otherwise measuring a common exonic variant in cis with the variant under assessment. Another metric commonly used with RNA-seq is by calculating the relative expression of splicing events measured by percent splicing index (PSI).^50^ The PSI value is defined by the number of reads supporting the alternative splicing event (i.e., the non-canonical/abnormal event) divided by the number of all reads in the region covering the splicing event. As noted above, it is important to consider factors such as the gene- and tissue-specific nature of the splicing result, experimental approach (e.g. use of nonsense mediated decay inhibitors, potential for PCR bias), and technology used to assay splicing. It is therefore important to calibrate thresholds for specific RNA sources and assay protocols, using known Pathogenic and Benign variants, to interpret variants as spliceogenic or non-spliceogenic and to consider their value for providing quantitative information. Further, combined evidence across studies of different design can provide complementary information and more confidence in result interpretation e.g. a patient-derived result without allele-specific quantification with accompanying mini-gene result providing evidence of the complete impact leading to the same splicing aberration.

In addition to the considerations highlighted above, another important aspect for result interpretation and weighting is how to assign PVS1/BP7 evidence strength to experimental data showing complex read-outs (two or more alternative transcripts, from the same allele). A logical approach is to assign a PVS1 evidence strength to each individual transcript, pool together transcripts with same evidence strength, and then apply an appropriately conservative “overall” evidence strength that considers the relative contribution of different transcripts to the overall expression (including full-length transcript).

### Application of PP3 and BP4 predictive codes for impact at the RNA splicing level

Computational splicing prediction tools provide key information for variant spliceogenicity (impact on splicing profile) and related pathogenicity (association with disease risk). To date, numerous splicing prediction tools have been used to inform application of computational codes PP3 and BP4 (and the dependent BP7 code). For examples see Table S3. However, there has been no standardised approach for deriving and applying prediction score thresholds for each tool. Here we show several examples of approaches to compare sensitivity and specificity of different prediction tools and how to apply computational codes based on tool outputs. Consistent with ClinGen recommendations for tool use by Variant Curation Expert Panels, we only assessed tools that were publicly available.

#### Comparison of splicing prediction tools

Eleven splicing prediction tools covering a wide variety of algorithmic approaches (Table S3) were compared to functional data from a saturation genome editing assay of 13 *BRCA1* exons^12^ that measured cell survival to infer whether splicing variants are Benign or Pathogenic. Importantly, most variants in this dataset are not present in repositories such as ClinVar or the Human Gene Mutation Database (http://www.hgmd.cf.ac.uk/ac/index.php), which several tools use in training their model. The ROC analysis of each splicing tool demonstrated that SpliceAI outperformed all other methods when assessing the area under the curve (AUC=0.959) for variants located at conserved positions at the donor and acceptor regions (Figure S5). These results are consistent with those reported in other publications.^6; 10; 51^

The strength of evidence associated with SpliceAI prediction score categories was also assessed using the same cell survival dataset.^12^ As shown in Table S9, SpliceAI score ≥0.5 (the cutoff recommended in the original SpliceAI paper ^12^) yielded strong evidence for variant impact on cell survival (i.e. inferred pathogenic; lower 95% confidence interval [CI] LR=18.58), while SpliceAI score <0.1 provided moderate evidence for no variant impact on cell survival (i.e. inferred Benign, lower 95% CI LR=0.03). Only a supporting level of evidence is required for application of PP3, but these data suggest that SpliceAI score ≥0.5 may be calibrated too high, leading to the exclusion of many spliceogenic variants from receiving a predictive code. Notably, this assessment used a single preselected SpliceAI cut point and inferred pathogenicity based on analysis of Multiplexed Assays of Variant Effect (MAVE) data and a relatively low number of variants (414 total) from selected functional domains of a single gene.

#### Model for establishing thresholds for splicing prediction tools

We demonstrate below a process for calibrating thresholds for different computational tools using large variant datasets with splicing assay results (Figure 2A). We investigated the optimal threshold values for SpliceAI (a tool trained on predicting splicing events, not pathogenicity) by comparing the Δ score output for 2736 variants located outside canonical splice sites across eight genes (*BRCA1, BRCA2, MLH1, MSH2, MSH6*, *PMS2, NF1,* and *POU1F1*), and with associated in vitro splicing assay data curated from the literature. Output from our web tool (https://gwiggins.shinyapps.io/lr_shiny/) was used to compare the sensitivity and specificity of different SpliceAI cut-points across three categories (see methods). This approach intentionally designated a middle category to represent a range of uninformative spliceogenicity scores and for which bioinformatic codes should not be applied (i.e. both PP3 and BP4 are not met). Based on this analysis, the optimal threshold for assigning PP3 to non-canonical splice site variants was determined to be ≥0.2, which equated to a moderate level of evidence for spliceogenicity (Table 2, Figure 2B, Figure 3). For all non-canonical splice site variants, this threshold provided 78% sensitivity for true spliceogenic variants. Sensitivity increased to 91% for the subset of non-canonical splice site variants located within the standard splice region (Donor site: last 3 bases of the exon and 6 nucleotides downstream from the exon boundary; Acceptor site: first base of the exon and 20 nucleotides upstream from the exon boundary) (Table S10). The optimal threshold for assigning BP4 was determined to be ≤0.1 for moderate level of evidence for non-spliceogenicity with 87% specificity for all non-canonical splice site variants and 73% specificity for variants within the standard splice region (Table 2). LRs for narrower bands of scores around the selected cutpoints were consistent with moderate evidence against spliceogenicity (e.g. SpliceAI Score range 0.08-0.1, LR=0.122) and supporting evidence for spliceogenicity (e.g. Score range 0.20-0.22, LR =2.52).

**Figure 2.**
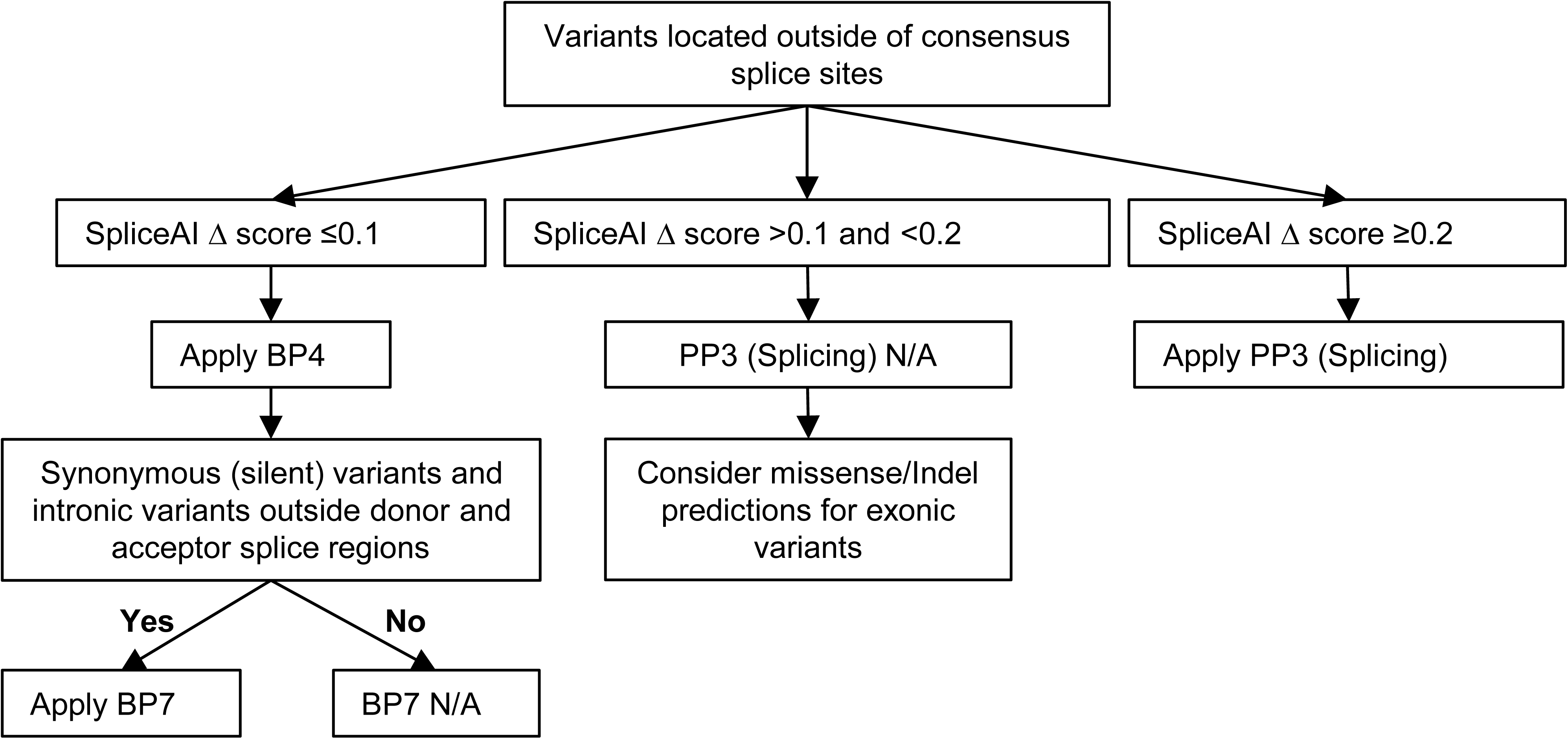
Model for optimising thresholds for prediction algorithms of alternative splicing. (A) Schematic demonstrating how collation of three variant datasets (in vitro splicing data, splicing prediction scores, and clinical classification data) enables calibration of splicing prediction algorithms for pathogenicity. While clinically classified variant data is preferable, splicing assay data can be used as an imperfect surrogate for pathogenicity. More extensive annotation of alternative splicing events and level of aberration will lead to an improved correlation of splicing events with variant pathogenicity. The distribution of hypothetical computationally predicted splice scores is illustrated, showing significant overlap of non-spliceogenic/spliceogenic datasets (left side) and Benign/Pathogenic datasets (right side). The low, intermediate and high prediction score used to assign ACMG/AMP code weighting can be determined by calculating likelihood ratios for different score categories, and obtaining consensus on the score thresholds to be applied. (B) Process for calibrating splicing prediction score thresholds for computational tools. A worked example of a likelihood ratio calculation is shown in Table S4. Note: truth datasets exclude splice site variants, which are captured by the PVS1 decision tree process.

**Figure 3.**
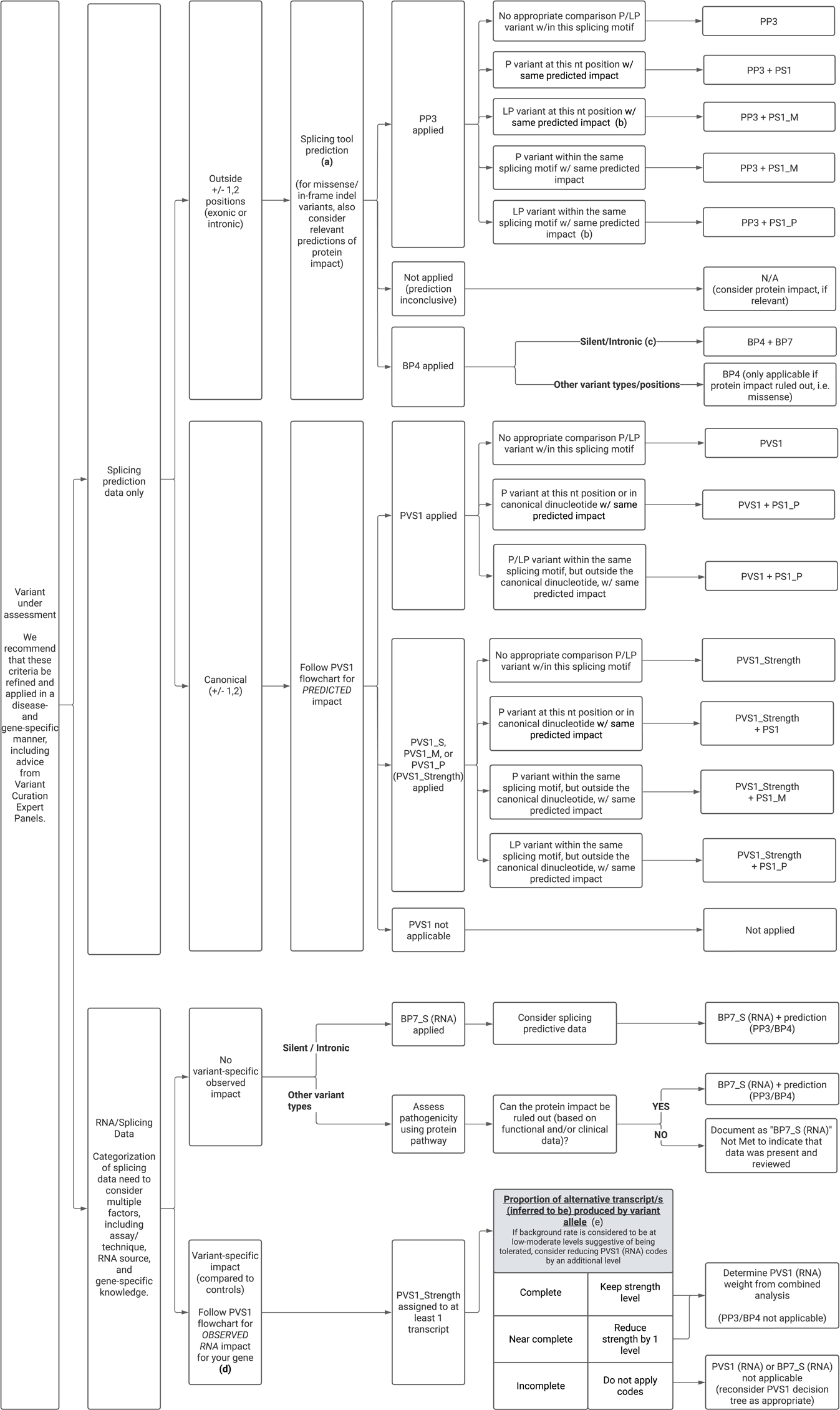
An exemplar PP3, BP4, and BP7 decision tree for maximum SpliceAI splicing prediction scores and calibrated cut-off scores. The analytical process is shown in Figure 2B and data shown in Table 2. BP7 should not be applied for splice motif regions given their higher prior. This may be defined as the standard splice region (a conservative application already implemented by several VCEPs) or the minimal splice region. PP3 may still be applied for missense or insertion-deletion variants that show computational evidence for a deleterious effect for change in protein sequence.

**Table 2.**
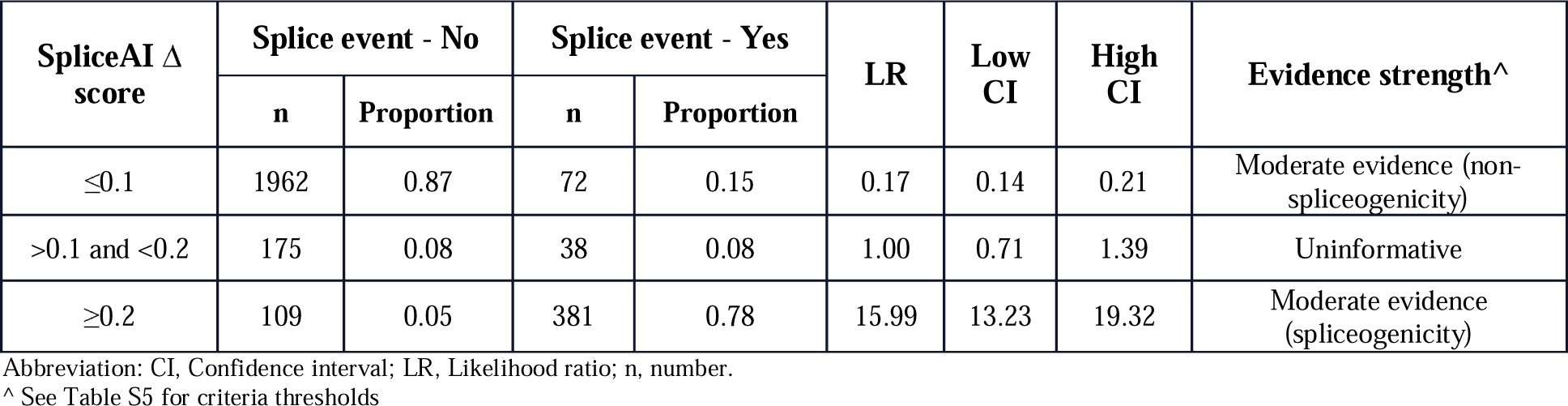
Likelihood ratio analysis of the maximum SpliceAI Δ score for non-canonical splice site variants using optimal cut-offs.

| SpliceAI $\Delta$ score | Splice event - No | | Splice event - Yes | | LR | Low CI | High CI | Evidence strength<sup>^</sup> |
| --- | --- | --- | --- | --- | --- | --- | --- | --- |
|  | n | Proportion | n | Proportion |  |  |  |  |
| $\leq 0.1$ | 1962 | 0.87 | 72 | 0.15 | 0.17 | 0.14 | 0.21 | Moderate evidence (non-spliceogenicity) |
| $>0.1$ and $<0.2$ | 175 | 0.08 | 38 | 0.08 | 1.00 | 0.71 | 1.39 | Uninformative |
| $\geq 0.2$ | 109 | 0.05 | 381 | 0.78 | 15.99 | 13.23 | 19.32 | Moderate evidence (spliceogenicity) |

Abbreviation: CI, Confidence interval; LR, Likelihood ratio; n, number.
<sup>^</sup> See Table S5 for criteria thresholds

It should be noted that the SpliceAI calibration results were not materially different when excluding large-scale construct-based datasets (data not shown). Although the results are based on a limited number of genes (given the availability of highly curated information on variant-associated splicing impact), they are expected to be widely applicable since the mechanism of RNA splicing is not gene-specific. It is notable that the threshold of 0.2 for predicting spliceogenicity is consistent with that defined by the developers of SpliceAI for a high recall threshold.^16^

The Moderate strength of evidence for spliceogenicity associated with the upper (≥0.2) and lower (≤0.1) SpliceAI thresholds did not change when assessing variants within the standard splice site motifs, and was at least Supporting for variants (exonic or intronic) outside the standard splice region (Table S10). Compared to the standard splice region, results from analysis restricting to variants within the minimal splice region (Table S11), showed similar positive predictions (92%) but more false positive predictions of spliceogenicity (28%). Additional analysis separating exonic and intronic variants outside the minimal splice region (Table S11) showed that positive prediction is much lower for exonic variants (58%, presumably due to poorer prediction of variants impacting ESEs), but negative prediction is good (84%). Both positive and negative predictions were good for intronic variants (>84%).

These observations are consistent with overall higher prior for spliceogenicity for variants located at highly conserved splice motif positions, and justify the conservative approach that BP7 should not be applied for variants within a designated splice region (standard or minimal). Findings also justify that application of BP7 is warranted for both synonymous exonic and intronic variants outside designated splice regions and with no predicted splice impact (BP4 met).

We recognise that not all spliceogenic variants will be Pathogenic - either due to incomplete effect (level of aberration) and/or type of resulting aberration/s (in-frame or rescue transcripts); however, at this point in time, current tools are not yet sufficiently developed to accurately predict these relationships. Further, there are many reasons why a specific aberration type may not be captured by existing tools (e.g. large intron retention, or multi-exon skipping). For this reason, combined with the results for sensitivity analysis considering LRs for narrow bands around selected cutpoints, a conservative approach is to calibrate codes to reach a moderate level of evidence, but only apply predictive PP3/BP4 codes at a supporting weight.

We provide a model for evaluating and calibrating individual bioinformatic tools. We demonstrate that prediction using a single tool, trained in a data-driven way to detect splicing impact via multiple mechanisms, can be sufficient to provide at least supporting level of evidence for application of BP4 or PP3. Future analysis with much larger datasets that allow more extensive stratification by variant location will provide more clarity on the applicability of higher weights, as has recently been recommended in the context of missense prediction.^52^ There will be continuing need to train, test and calibrate tools to improve prediction for variants resulting in pseudoexonization (influenced by a range of factors) and/or variants that impact atypical (non-GT-AG) splice sites or splicing elements with relatively poor conservation and/or redundancy such as exonic and intronic splicing enhancers/silencers and branchpoints.^10; 53–55^ Where a single tool cannot provide adequate prediction across multiple mechanisms, there may be value in using complementary tools to detect splicing aberrations due to different mechanisms following a decision-tree process.^56^ Where possible, gene-specific knowledge (e.g. naturally-occurring isoform patterns) should be considered in assessing and applying bioinformatic scores for predicted splicing as part of variant curation protocols.

### Application of the BP7 computational code

The description for BP7, as drawn from the original ACMG/AMP publication,^1^ is as follows: A synonymous (silent) variant for which splicing prediction algorithms predict neither an impact to the splice canonical sequence nor the creation of a new splice site AND the nucleotide is not highly conserved. Review of existing VCEP definitions for this code indicate variability in application, particularly in relation to consideration of conservation and relevance to intronic variation. We highlight the following issues to consider regarding use of this code to capture bioinformatic predictions.

First, Richards et al., 2015^1^ recommend that BP7 is applicable *after assignment of BP4 for no adverse splicing predictions* in order to capture the low prior probability of pathogenicity of silent variants. We further recommend that BP7 should not be applied for synonymous substitutions in certain locations, specifically those located at the first base or the last three bases of the exon, with higher likelihood to impact splicing due to disruption of the acceptor or donor motif. Motif range may be altered as appropriate for non-GT-AG introns.

Second, we caution against inclusion of evolutionary conservation in assessment of silent or intronic variants for application of code BP7, without empirically-derived justification for choice of tool, evolutionary depth, and cutpoint to define a nucleotide as “not highly conserved”. Published evidence from an analysis of 27,733 variants within or adjacent to 2,198 human exons shows that while mean conservation score is higher for splice-disrupting variants (as expected given the nucleotide conservation is a feature of functional motifs), conservation has limited predictive power to detect non-splice disrupting variants ^47^. Further, secondary analysis of our curated dataset indicates that, for silent and intronic variants for which BP4 might be applied on the basis of SAI score ≤0.1, fewer variants would have BP7 applied with no improvement in negative predictive value for applying an additional filter of “not highly conserved” (Table S12). This likely reflects that splicing prediction tools implicitly capture conservation in the context of predicted variant impact via abrogation of function of splicing motifs, while position weight matrices also allow for some nucleotide sequence variation within the motif.

Third, we acknowledge and concur with protocols for many diagnostic testing laboratories and VCEPs that the BP7 code may be applied for intronic variants, assuming that mechanism of possible effects will be on mRNA processing, as invoked for silent variants. Currently 10 VCEPs (Brain Malformations, CDH1, DICER and miRNA-Processing Gene, ENIGMA BRCA1 and BRCA2, Glaucoma, Hereditary Breast Ovarian and Pancreatic Cancer, InSiGHT Hereditary Colorectal Cancer/Polyposis, Myeloid Malignancy, PTEN, and RASopathy) refer to the use of BP7 to include intronic variant classification (Table S1). Guidelines from six VCEPs (CDH1, DICER and miRNA-Processing Gene, ENIGMA, Hereditary Breast Ovarian and Pancreatic Cancer (*PALB2*), InSiGHT Hereditary Colorectal Cancer/Polyposis (*APC*), and PTEN) specifically note that BP7 is only applicable for intronic/non-coding variants at or beyond positions +7/-21 with no predicted effect on splicing and that this code is applied in addition to BP4. The Hereditary Breast Ovarian and Pancreatic Cancer VCEP has designated BP7 application for variants at or beyond positions +7/-40 in the *ATM* gene. The ENIGMA BRCA1 and BRCA2 VCEP specifications justify the application of BP7 for variants outside splice motifs based on maximum likelihood estimation analysis of breast cancer case-control data^57^ with results indicating that location of an intronic variant at or beyond positions +7/-21 provides moderate evidence against pathogenicity - even without applying a bioinformatic prediction filter.

Reassessment of the designation of intronic nucleotide boundaries and the strength of evidence for BP7 code application (in addition to BP4) for intronic variants over time will be important as more information accrues from sequencing of clinical cohorts. Until such time, we consider a conservative application of BP7 as a predictive computational code for intronic variants located outside the standard splice region (at or beyond positions +7/-21) having also met BP4, consistent with application by several VCEPs to date (as noted above). However, we acknowledge, based on the analysis presented here (Table S11), there is rationale to expand application of BP7 to intronic variants located outside the minimal splice region. [Note, this restriction with respect to intronic variant location is not relevant to the application of BP7_Strong to capture RNA results demonstrating no impact on splicing].

### Adaptation of the PS1 code to incorporate splicing predictions

The ACMG-AMP code PS1 is strong pathogenic evidence defined as “*same amino acid change as a previously established Pathogenic variant regardless of nucleotide change*”. The underlying premise for the PS1 code is that the clinical evidence for a previously classified missense variant can be applied to infer pathogenicity for the same predicted molecular change at the protein level. Although both the Pathogenic variant and variant under assessment (VUA) should have no predicted/confirmed effect on RNA splicing for PS1 to be applied for a missense variant, there is <u>no specific requirement for experimental validation</u> of the predicted molecular effect of the amino acid change encoded by the Pathogenic variant or the VUA.

There is thus rationale to apply code PS1 in the context of exonic and intronic variants based on similarity of predicted RNA effects for a VUA in comparison to a known Pathogenic variant. This follows the logic that this PS1 code captures existing evidence of pathogenicity for a variant with an identical mechanism of pathogenicity. Indeed, this concept has been used in specifications (or pilot specifications) for four different ClinGen Variant Curation Expert Panels (Hearing Loss, ENIGMA BRCA1 and BRCA2, Hereditary Breast Ovarian and Pancreatic Cancer, and *PTEN*), and also by the Cancer Variant Interpretation Group UK (CanVIG) (Table S1). However, the recommendations differ between four of the five expert groups in relation to the relative location of the two nucleotide changes, applicability to variants at the canonical splice dinucleotide positions, applicability to exonic variants, code weight, and/or applicability when the previously classified variant has Likely Pathogenic assertion versus Pathogenic assertion. Further, the current specifications for the MMR gene VCEP denote ACMG-AMP code PM5 (originally defined as “*missense change at an amino acid residue where a different missense change determined to be Pathogenic has been seen before*”) in the context of predicted splicing for a variant under assessment, compared to both prediction and splice assay data for a comparison variant, namely “*Variants affecting the same splice site as a confirmed splice variant with similar or worse splicing in silico prediction*s”.

Here we provide more guidance on the application of a PS1 code with justification for differences in weights considering the relative locations of the variant being assessed and the previously classified (Likely) Pathogenic variant (Table 3). The concept of PM5 for splicing prediction is not considered directly applicable, since the recommendation is to apply PS1 only when the predicted event of the known (Likely) Pathogenic variant precisely matches the predicted event of VUA e.g. where the effect is via impact on the same motif, such as two different variants within the same donor splice site motif that are both predicted with similar scores to disrupt function of that donor site, or two different variants in the same splice region that are both predicted with similar scores to lead to use of the same cryptic site.

**Table 3.** PS1 code weights for variants with same predicted splicing event as a known (Likely) Pathogenic variant*

| Variant Under Assessment (VUA) | Baseline computational/predictive code applicable to VUA | Position of comparison variant relative to VUA | PS1 code applicable to VUA |  |
| --- | --- | --- | --- | --- |
|  |  |  | with P comparison variant | with LP comparison variant |
| Outside canonical dinucleotide | PP3 | Same nucleotide | PS1 | PS1_Moderate |
| Outside canonical dinucleotide | PP3 | Within same splice motif (including within canonical dinucleotide) | PS1_Moderate | PS1_Supporting |
| Canonical dinucleotide | PVS1 | Within same canonical dinucleotide | PS1_Supporting | N/A |
| Canonical dinucleotide | PVS1 | Within same splice motif, but outside canonical dinucleotide# | PS1_Supporting | PS1_Supporting |
| Canonical dinucleotide | PVS1_Strong, PVS1_Moderate, or PVS1_Supporting | Within same canonical dinucleotide | PS1 | N/A |
| Canonical dinucleotide | PVS1_Strong, PVS1_Moderate, or PVS1_Supporting | Within same splice motif, but outside canonical dinucleotide# | PS1_Moderate | PS1_Supporting |

\* Prerequisite for all: The predicted event of the VUA must precisely match the predicted event of the comparison (Likely) Pathogenic variant (e.g. both predicted to lead to exon A skipping, or both to enhanced use of cryptic site B) AND the strength of the prediction for the VUA must be of similar or higher strength than the strength of the prediction for the comparison (Likely) Pathogenic variant. For an exonic variant, predicted or proven functional effect of missense substitution/s encoded by the VUA and (Likely) Pathogenic variant should also be considered before application of this code. Canonical dinucleotide refers to donor and acceptor dinucleotides in reference transcript/s used for curation. Designated donor and acceptor site motifs ranges should be based on position weight matrices for intron category (see methods). For GT-AG introns these are defined as follows: the donor site motif, last 3 bases of the exon and 6 nucleotides of intronic sequence adjacent to the exon; acceptor site motif, first base of the exon and 20 nucleotides upstream from the exon boundary. Consider other motif ranges for non-GT-AG introns.

**Table 4.** ACMG/AMP codes recommended for recording evidence relevant to variant position, and predicted and experimentally observed impact on splicing

| ACMG/AMP Code | Original definition<br>(Richards et al 2015) | (Re)definition for RNA impact | Notes |
| --- | --- | --- | --- |
| PVS1 | PVS1. Null variant (nonsense, frameshift, canonical +/-1 or 2 splice sites, initiation codon, single or multi-exon deletion) in a gene where loss of function is a known mechanism of disease. | *Bioinformatic data only* - Canonical splice site variants ($\pm 1,2$) in a gene with established LOF as a disease mechanism. | Develop gene-specific decision tree where feasible.<br>Use PVS1 decision tree to determine code strength. |
| PVS1_Strength (RNA) | - | *Splicing assay data* - Assays demonstrating a variant leads to aberrant splicing profile that can be categorised against a PVS1 decision tree. | PVS1_Strength (RNA) to be used to designate capture of splicing data (not PS3).<br>Use PVS1 decision tree to determine code strength. See Figure 4 flowchart. PVS1_Strength (RNA) may not be applicable for variants for which there is a plausible rescue model |
| PS1 | PS1. Same amino acid change as a previously established Pathogenic variant regardless of nucleotide change. | Same predicted splicing impact as a previously classified (Likely) Pathogenic variant. | <u>Predicted event</u> of variant matches that of a known (Likely) Pathogenic variant.<br>Weights depend on relative positions of the variant under assessment and confidence in classification for the comparison variant. See Table 3. |
| PS3 | PS3. Well-established in vitro or in vivo functional studies supportive of a damaging effect on the gene or gene product. | Not applicable for splicing effects. | Replaced by PVS1_Strength (RNA), with weight determined as per PVS1 decision tree and other factors. See Figure 4 flowchart. |
| PP3 | PP3. Multiple lines of computational evidence support a deleterious effect on the gene or gene product (conservation, evolutionary, splicing impact, etc.).<br>*Caveat: Because many in silico algorithms use the same or very similar input for their predictions, each algorithm should not be counted as an independent criterion. PP3 can be used only once in any evaluation of a variant.* | Computational evidence from calibrated prediction tool/s supports impact on splicing. | No requirement for multiple tools, but calibration of tool/s to select cutoffs for predicting spliceogenicity is recommended (gene-specific calibration not required).<br>Only use for non-canonical splice site variants (outside of $\pm 1,2$) when splicing assay data is not available.<br>For exonic variants, also consider functional impact via encoded change in protein sequence. |
| BS3 | BS3. Well-established in vitro or in vivo functional | Not applicable for splicing effects. | Replaced by BP7_Strong (RNA). See Figure 4 |
|  | studies supportive of a damaging effect on the gene or gene product. |  | flowchart. |
| BP4 | BP4. Multiple lines of computational evidence suggest no impact on gene or gene product (conservation, evolutionary, splicing impact, etc.)<br>*Caveat: Because many in silico algorithms use the same or very similar input for their predictions, each algorithm cannot be counted as an independent criterion. BP4 can be used only once in any evaluation of a variant.* | Computational evidence from calibrated prediction tool/s supports no impact on splicing | • No requirement for multiple tools, but calibration of tool/s to select cutoffs for predicting spliceogenicity is recommended (gene-specific calibration not required).<br>• May be applied in conjunction with BP7 and its derivatives for intronic/silent variants. See Figure 4 flowchart.<br>• For exonic variants, also consider functional impact via encoded change in protein sequence. |
| BP7 | BP7. A synonymous (silent) variant for which splicing prediction algorithms predict no impact to the splice consensus sequence nor the creation of a new splice site AND the nucleotide is not highly conserved. | Synonymous (silent) variant OR intronic variant with low prior probability of pathogenicity if no predicted impact on splicing. | • Apply only if BP4 is met.<br>• Evolutionary conservation is not considered informative for application of this code.<br>• Only applicable for intronic/non-coding variants outside the designated splice region (conservatively at or beyond positions +7/-21 and synonymous (silent) exonic variants located outside of the first and the last 3 bases of the exon, otherwise minimal splice region). |
| BP7_Strong (RNA) | - | Splicing assay data demonstrating a variant is not associated with aberrantly spliced transcript/s relative to transcript profiles in controls. | • BP7_Strong (RNA) to be used to designate capture of splicing data (not BS3).<br>• See Figure 4 flowchart for guidance on weighting and combining with other codes.<br>• For exonic variants, also consider functional impact via encoded change in protein sequence. |

Application of the code varies depending on whether the VUA and the comparison known (Likely) Pathogenic variant are located outside or inside a canonical splice site (see Table 3), and VUA location relative to the (Likely) Pathogenic variant. PS1 is applied at full strength for a variant located outside the canonical dinucleotide with a predicted RNA event that matches (with similar or higher strength of prediction) that of another Pathogenic variant at the same nucleotide. PS1 may be applied at moderate weight for a VUA in the event that the comparison variant is classified as Likely Pathogenic under the assumption that some clinical information would have been required to apply this classification for a non-canonical splice site variant. The predicted splicing event could be the result of any combination of loss of the native splice sites or gain/strengthened splice sites, but as noted above, the predicted event for the known (Likely) Pathogenic variant must match the predicted event of the VUA.

There is also allowance for comparison of a VUA to a (Likely) Pathogenic variant at different nucleotide positions outside the canonical splice site. We recommend limiting the comparisons to variants located within the same core splice donor or acceptor motif and reducing criterion weight by one strength level. These restrictions recognise the chance that, despite similarity in predicted impact, the variant position may possibly be associated with differences in type/s and level of RNA aberration/s produced. We suggest that the designation of donor and acceptor site motifs be based on position weight matrices (see footnote to Table 3 for more information).

PS1 may also be applied for a variant located at the canonical splice dinucleotides and for which there is another (Likely) Pathogenic variant located within the same splice motif (including the canonical splice site). However, in most instances the proposed strength levels are reduced to prevent overweighting of the VUA compared to the original (Likely) Pathogenic comparison variant. Suggested weights consider whether the comparison variant lies within the same canonical dinucleotide or at other positions within the same motif, the PVS1 weight applicable to the VUA, and whether the comparison variant is classified as Pathogenic or Likely Pathogenic.

### Combined application of ACMG/AMP codes that capture evidence relating to variant location, splicing predictions, splicing assay data, and variant type

As denoted, multiple ACMG/AMP criteria may be suitable to record structured evidence relevant to a variant’s (predicted) impact on splicing. We propose re-purposing of existing evidence codes (PVS1, BP7) to capture mRNA splicing assay data separately from other protein or cell-based functional assay data (recorded under PS3/BS3), and abrogating bioinformatic prediction codes when splicing assay data is used to designate a variant as loss-of-function following the PVS1 decision process.

Figure 4 provides a general scheme of code application and strength based on variant position, bioinformatic predictions, other (Likely) Pathogenic variants with similar predicted impact on splicing, and how to add, replace or modify codes based on RNA/splicing assay data. We anticipate that the more challenging aspects to implementing these splicing interpretation recommendations will be: (i) decisions on similarity in predicted effect when comparing predictions for a VUA against a known (Likely) Pathogenic variant; (ii) assigning baseline PVS1 code weights; (iii) consideration of factors that are relevant to determining and/or downweighting of PVS1 or BP7 codes based on splice assay results, including partial splicing impact or complex aberration profiles; (iv) removal of PP3/BP4 prediction codes upon upgrading of a variant to PVS1 on the basis of splicing data (consistent with recommendations to not use PP3 together with PVS1); (v) recording - but not applying a code for - splicing data for missense or in-frame insertion/deletion variants in the absence of protein functional data that is required to fully account for all potential mechanism/s of variant impact.

**Figure 4.**
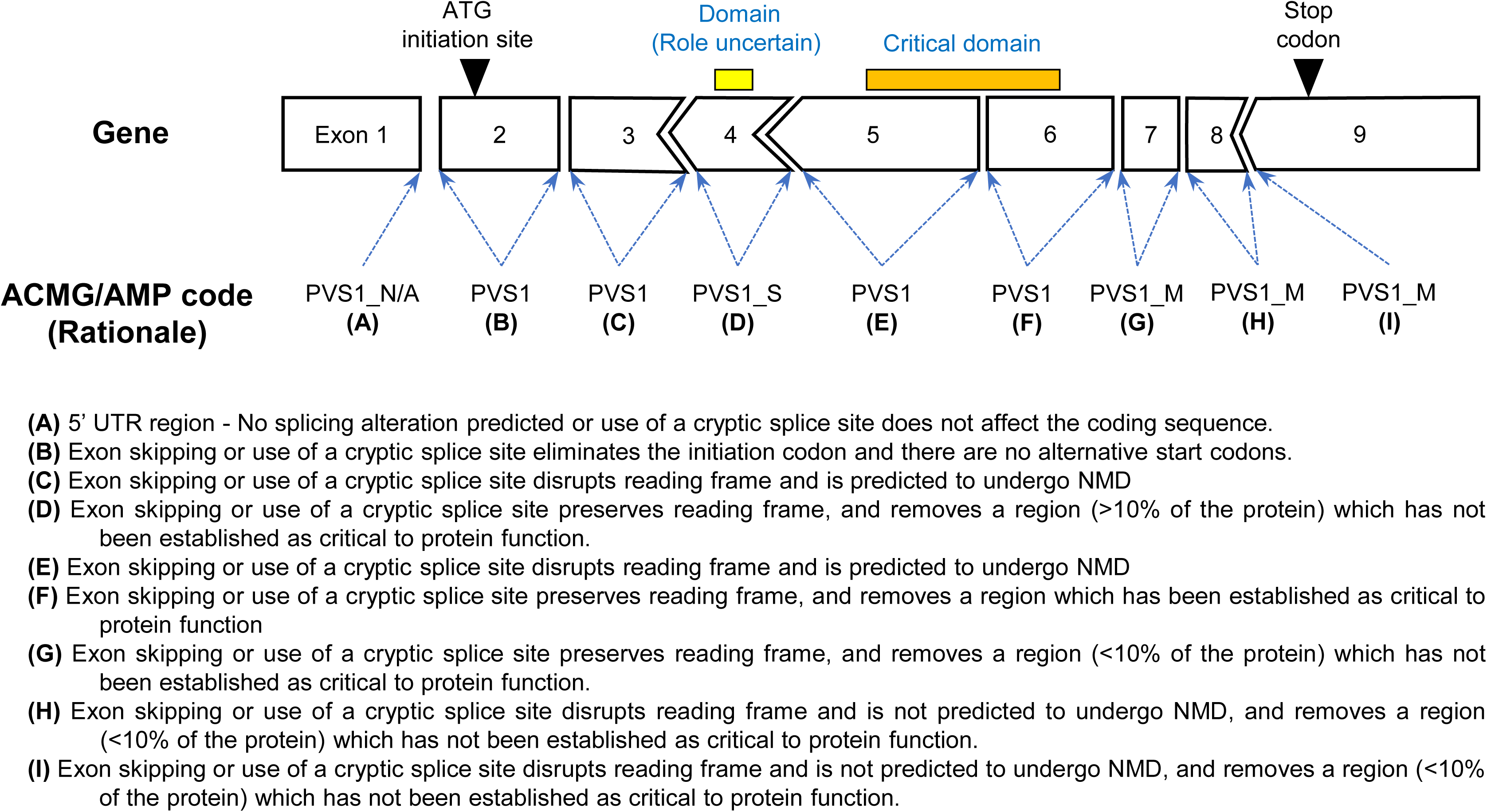
Decision tree for application of bioinformatic codes and RNA splicing assay results for variant interpretation. Footnotes: (a) Alternative prediction tools/thresholds may be appropriate for variants that impact sites other than GT-AT donor-acceptor motifs. (b) LP variants at the canonical positions should only be used as evidence if additional supporting clinical evidence is present. (c) Silent (excluding last 3nt of exon and first nt of exon) and intronic variants at or beyond the +7 and −21 positions (conservative designation for splice region) or otherwise as at or beyond the +7 and −4 positions (less conservative designation for minimal splice region). (d) If multiple impacts are observed from a splicing assay, use flowchart for the most conservative application of PVS1 based on experimental data. (e) We recommend that these thresholds be refined and applied in a disease- and gene-specific manner, including advice from Variant Curation Expert Panels. Categorization as complete or near complete needs to consider multiple factors, including assay/technique, RNA source, and validation of assay weights using established controls. Examples of laboratory-specific approaches and suggested operational thresholds have been reported previously.^31; 58–61^

Accounting for all potential disease mechanisms of other coding variant types (e.g. missense, in-frame insertions/deletions) can be exceptionally challenging during variant classification. We generally recommend that the effects on the protein with and without splicing impact be evaluated independently as separate variant classifications based on these different mechanisms, with the most deleterious classification resulting in the final classification. For variants with experimental splicing assay data indicating no impact, we conservatively recommend retaining the most deleterious effect of the bioinformatic prediction (i.e. the most severe impact of splicing or other variant effect, such as a missense alteration). Splicing results should be recorded as explanatory text but BP7_Strong should not be applied in the final classification of such coding variants. In cases where experimental data indicates no functional impact for the other coding variant effects and experimental splicing assay data indicates no impact on splicing, then BP7_Strong may be applied to allow for optimal tracking of experimental data. Rarely, if relevant gene-specific considerations obviate need for functional assays for other coding variant effects, then BP7_Strong may also be applied.

## CONCLUSIONS

This manuscript provides recommendations regarding use of ACMG/AMP codes to better categorize splicing prediction and/or laboratory (splicing assay) evidence, including clarification on the application of existing splicing-related codes and re-purposing of other codes to capture splicing-related evidence. We describe generic protocols useful for assigning strength levels to different evidence criteria, to facilitate recalibration of code strengths as new information accrues. We also provide a generic decision-tree to guide variant assessment with combinations of evidence codes relating to variant location, splicing predictions, splicing assay data and variant type.

The framework presented herein is generally applicable to the majority of canonical GT-AG splicing consensus sites observed in >98% of splice sites in mammalian genomes.^19^ Evaluation of other non-canonical dinucleotide pairs (e.g. GC-AG) used for splicing or variation in genomic regions with low prior probability of altering splicing (e.g. branch points, deep intronic leading to pseudoexonization) may also result in splicing aberrations. It is anticipated that these additional mechanisms underlying splicing aberrations will most likely be identified through the detailed evaluation of unexplained clinical cases through experimental RNA assay data rather than genomic DNA analysis. As more data accrues, it will be important to assess and further refine bioinformatic tool predictive capacity. Such efforts will benefit from large-scale high throughput studies that have not been directed towards clinically detected variants to mitigate against potential bias towards alterations at more conserved sites.

We provide a web-based tool that can be used to recalibrate using new datasets or subsets of datasets (https://gwiggins.shinyapps.io/lr_shiny/). To ensure gene- and disease-specification requirements may be assessed, we recommend that ClinGen VCEPs and other expert groups continually improve on these recommendations as their knowledge of clinically relevant transcripts and their variation in expression evolve through the use of new tools and molecular techniques.

## Data Availability

All data produced in the present work are contained in the manuscript

## ACKNOWLEDGMENTS

ABS and MTP were supported by Australian NHMRC Funding (APP177524). LCW and GARW were supported by Health Research Council funding (19/460 and 22/187) and the Mackenzie Charitable Foundation. We thank Hadley Northcott for his contribution to the development of the *BRCA1/BRCA2* splicing dataset. MdlH is supported by a grant from the Spanish Ministry of Science and Innovation, Plan Nacional de I+D+I 2013-2016, ISCIII (PI20/00110) co-funded by FEDER from Regional Development European Funds (European Union). ABB and SMH were supported by NIH grant U24 HG006834. The ClinGen Consortium Sequence Variant Interpretation Working Group members include Leslie G. Biesecker, Steven M. Harrison (co-chairs), Ahmad Abou Tayoun, Jonathan S. Berg, Steven E. Brenner, Alicia B. Byrne, Garry R. Cutting, Sian Ellard, Marc S. Greenblatt, Peter Kang, Izabela Karbassi, Rachel Karchin, Jessica Mester, Anne O’Donnell-Luria, Tina Pesaran, Sharon E. Plon, Heidi L. Rehm, Natasha T. Strande, Sean V. Tavtigian, and Scott Topper. ClinGen is primarily funded by the National Human Genome Research Institute (NHGRI) with co-funding from the National Cancer Institute (NCI), through the following grants: U24 HG009649 (to Baylor/Stanford), U24 HG006834 (to Broad/Geisinger), and U24 HG009650 (to UNC/Kaiser). The content is solely the responsibility of the authors and does not necessarily represent the official views of the National Institutes of Health.

## AUTHOR CONTRIBUTIONS

L.C.W., A.L., L.M.V., T.P., R.K., S.H. and A.B.S. conceived the study design. The complete writing group consisted of L.C.W., M.DLH., A.L., L.M.V., A.B.B., T.P., R.K., S.H. and A.B.S. A.L. provided a dataset comprising individuals from a clinical cohort. Data analysis was performed by L.C.W., G.A.R.W., A.L., M.T.P., D.M.C., D.B., A.C., A.T., and H.Z. S.H. chaired the ClinGen SVI Splicing Working Group and presented the data to the ClinGen SVI Working Group. All authors read and approved the final manuscript. The funders had no role in the design of the study, the collection, analysis, or interpretation of the data, the writing of the manuscript, or the decision to submit the manuscript for publication.

