## Supplemental Information for "APPLICATION OF THE ACMG/AMP FRAMEWORK TO CAPTURE EVIDENCE RELEVANT TO PREDICTED AND OBSERVED IMPACT ON SPLICING: RECOMMENDATIONS FROM THE CLINGEN SVI SPLICING SUBGROUP"

### Box S1. Biologically relevant transcripts and the rescue transcript model.

A rescue model was first proposed (referred to by the authors as reduced haplo-insufficiency) to explain attenuated familial adenomatous polyposis associated with a stop gain variant c.1192_1193del p.(Lys398Glufs*5) in the *APC* gene (Young et al., 1998). The stop-gain variant is located in an *APC* exon 9 region that undergoes physiological alternative splicing due to the presence of a strong internal acceptor. The resulting physiological alternatively spiced acceptor shift isoform r.934_1236del p.(Val312_Gln412del), also annotated as Δ(E9p303), splices out the stop-gain variant and, equally relevant, is expressed at high levels in normal colonic mucosa.

More recently, a similar rescue transcript mechanism has been proposed to explain the fact that *BRCA1* haplotype c.[594-2A>C;641A>G], causing exon 10 skipping (a stop-gain alteration), is not a high-risk variant (de la Hoya et al., 2016). In this case, the rescue mechanism was attributed to a physiological isoform splicing out exons 9 and 10. This Δ(E9,10) isoform accounts for 20-30% of the overall *BRCA1* expression level in most tested tissues, including normal breast and ovarian epithelia.

As expected, neither *APC* Δ(E9p303) nor *BRCA1* Δ(E9,10) target domains and/or residues presumed critical for protein function.

Figures S1-3 illustrate why it is critical to identify transcripts (in addition to the reference transcript) that code for a functional (or partially functional) protein, and to determine if the physiological expression level of each transcript is compatible with their potential to act as rescue transcripts, henceforth referred to as candidate rescue transcripts.

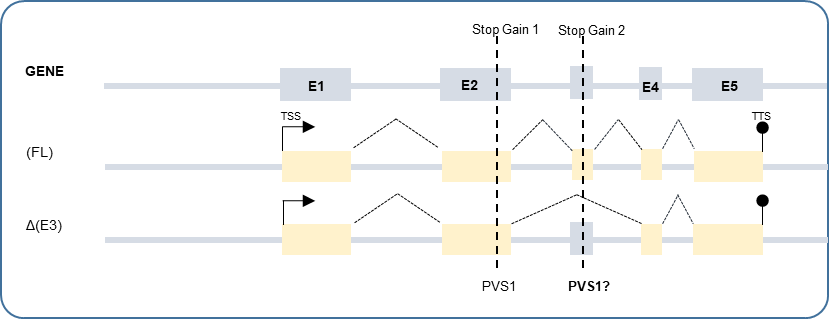

### Figure S1.

An example of candidate rescue alternative splicing. Assays targeting partial transcript sequences has identified an alternative exon 3 skipping event. Regardless of the biological function and/or relative expression level of these events, stop gain variant 1 (located in constitutive exon 2) is a *bona-fide* loss-of-function variant (predictive code PVS1). By contrast, stop gain 2 (located in exon 3) does not target exon 3 skipping transcripts, and is not necessarily a *bona-fide* loss-of-function variant. Ultimately, loss-of-function status will depend on the coding impact of exon 3 skipping, and its relative contribution to the overall gene expression. If coding for a (partially) functional protein, the Δ(E3) transcript might be a *bona-fide* rescue transcript, and stop gains located in exon 3 would not be associated with risk (or associated with lower risk than stop-gain variants in constitutive exon 2). Abbreviations: E, exon; FL, Full length; TSS, Transcription start site; TTS, Transcription termination site.

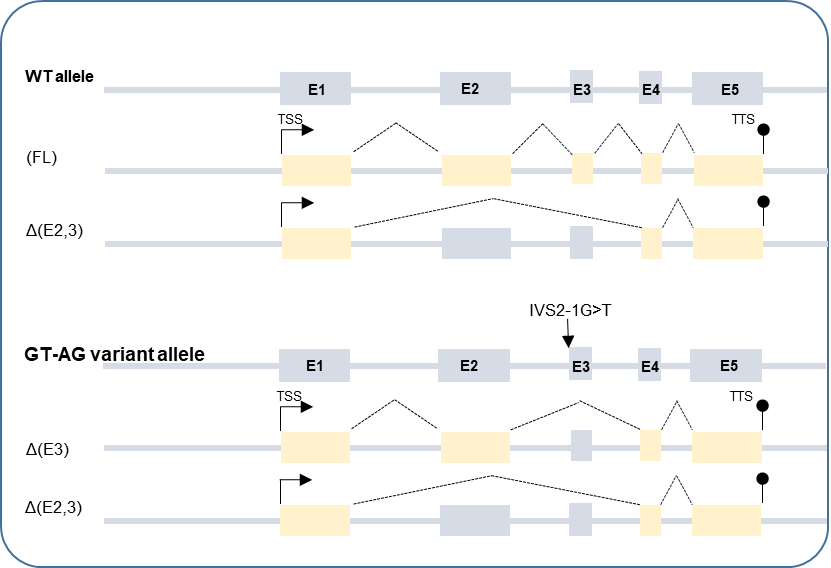

### Figure S2.

An example of a naturally occurring alternatively spliced transcript rescuing GT-AG splice site variants. In this example, the gene expresses two naturally occurring alternative splicing isoforms: full-length [FL] and Δ(E2,3). A genetic variant targeting the exon 3 acceptor site is predicted (or observed) to generate only Δ(E3) and Δ(E2,3) transcripts. If normal expression of Δ(E2,3) transcripts is able to provide sufficient functionality, IVS2-1G>T is not predicted to be a loss-of-function variant, even if exon 3 skipping is a PTC-NMD transcript. If this is the case, Δ(E2,3) will act as a naturally occurring rescue transcript for IVS2-1G>T. Furthermore, the Δ(E2,3) rescue transcript may be generated from any GT-AG variant targeting exons 2 and 3.

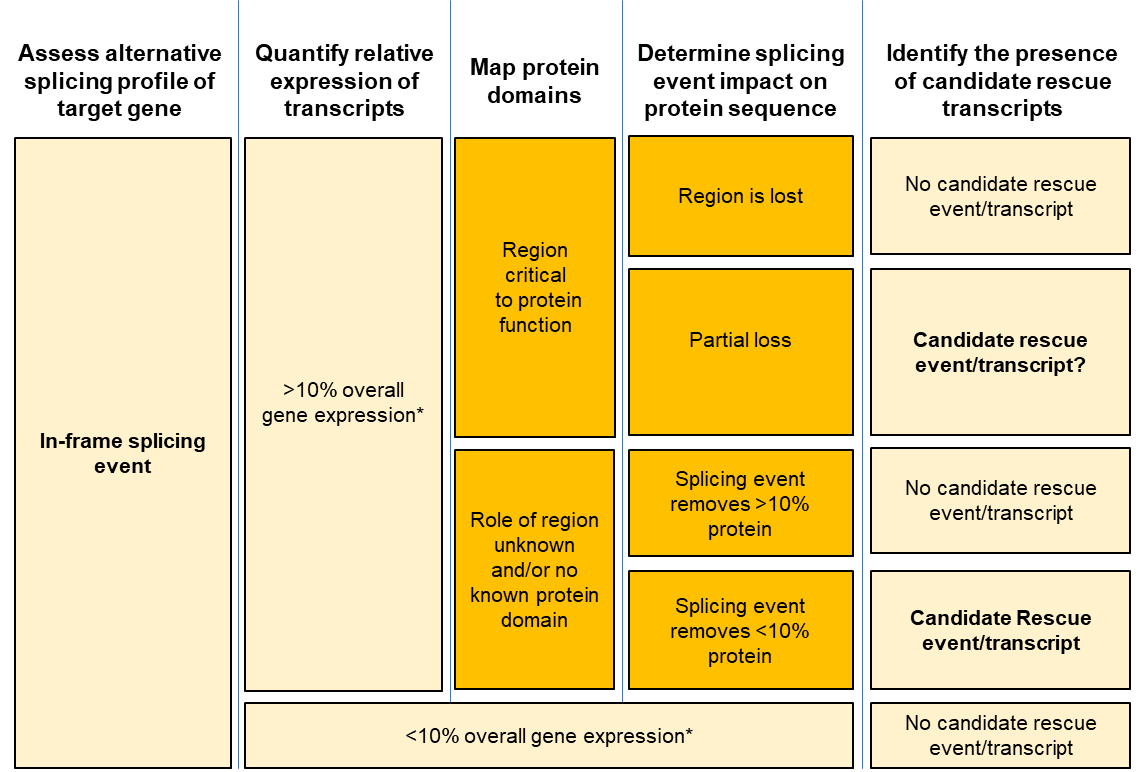

### Figure S3.

In-frame splicing event and the identification of candidate rescue transcripts. The figure highlights a gene-specific rationale to assess the likely functional impact of an in-frame alteration and its potential to act as a rescue transcript. After developing a gene-specific adaptation of the PVS1 decision tree, we propose using the same gene-specific rationale to annotate the physiological in-frame alternative splicing events as candidate rescue transcripts. The relative contribution of a naturally occurring in-frame event to the overall expression must first be considered when assessing the predicted functional impact. Secondly, the location of critical functional domain(s) is mapped relative to the in-frame event, to provide further guidance regarding potential for rescue. Consideration must also be given to the protein region lost if re-initiation occurs from the nearest in-frame ATG, and for stop gains at the 3’ end of the open reading frame. * Note that the threshold of 10% for defining potential rescue by an in-frame event (i.e., residual expression providing some degree of haplo-sufficiency) is operational. We recommend that this threshold be refined and applied in a disease- and gene-specific manner by Variant Curation Expert Panels.

A. Acceptor site variant.

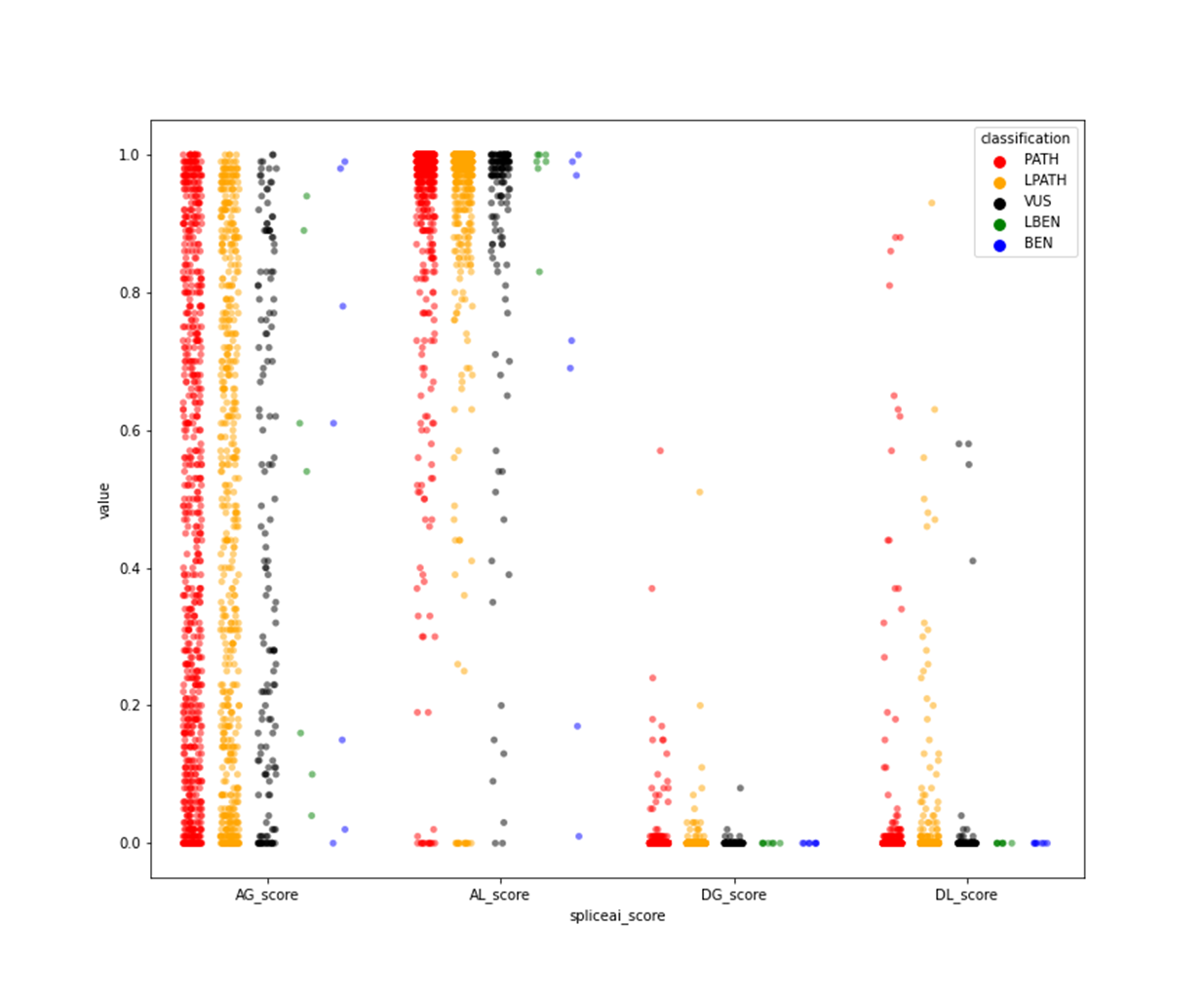

B. Donor site variants

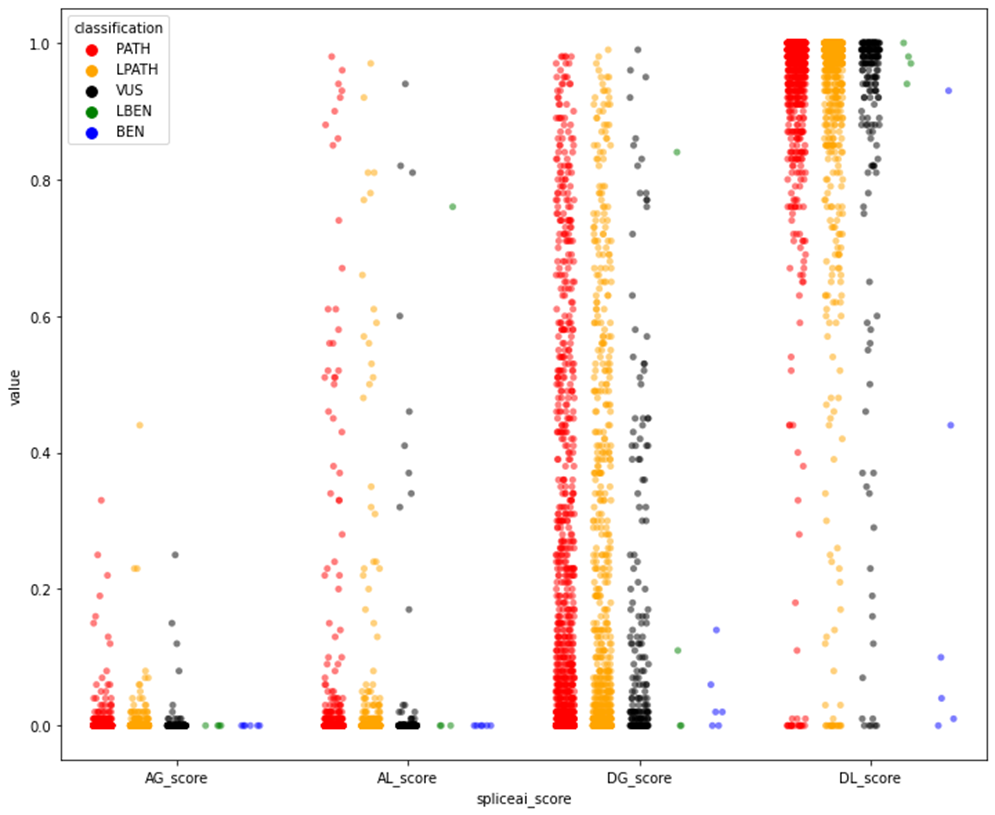

### Figure S4.

Dot plots summarising classifications and SpliceAI scores for canonical splice site (±1,2) variants found in diagnostic data from GeneDx. Information is included for 1043 genes designated as disease-causing through a loss-of-function mechanism. (A) Variants at acceptor sites (Total n=1418) (Likely/pathogenic, n=1254; VUS, n=151; Likely/benign, n=13), and (B) Variants at donor sites (Total n=1982) (Likely/pathogenic, n=1777; VUS, n=196; Likely/benign, n=9). Abbreviations: PATH=pathogenic; LPATH = likely pathogenic; VUS = variant of uncertain significance; LBEN = likely benign; BEN = benign; AG =Acceptor gain; AL = Acceptor loss; DG= donor gain; DL = donor loss. As expected for high probability of disruption to splicing for variants at native acceptor and donor sites, enrichment of high AL scores is seen for variants at acceptor sites (A), and enrichment of high DL scores is seen for variants at donor sites (B).

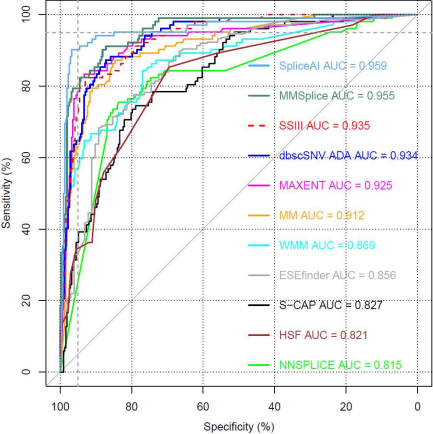

### Figure S5.

Receiver Operating Characteristic (ROC) curves of 11 splicing prediction tools using variants outside of the +/- 1/2 positions from the Findlay et al dataset (Findlay et al., 2018). Dataset included 414 variants at the donor and acceptor region (3 nucleotides synonymous and 8 nucleotides intronic).
